# Cognition, Lifestyles, and Environments: Quantifying the Roles of Body Physiology and the Brain

**DOI:** 10.64898/2026.02.26.26347222

**Authors:** Irina Buianova, Narun Pat

**Affiliations:** Department of Psychology, University of Otago, Dunedin, New Zealand

**Keywords:** Cognition, Aging, Machine Learning, Neuroimaging, Lifestyles

## Abstract

Lifestyle and environmental factors are increasingly promoted as modifiable targets for maintaining cognitive health. Yet, it remains unclear how well they predict cognitive functioning and to what extent their associations with cognition are reflected in brain and body measures. We applied machine learning to multimodal data from over 10,000 UK Biobank participants to evaluate the predictive value of twelve lifestyle and environment domains, spanning physical activity, diet, smoking, alcohol consumption, sleep, sexual behavior, electronic device use, and environmental exposures, for cognition, both individually and in combination, and quantified the extent to which these associations are captured by body and brain. A model integrating lifestyle and environment domains explained 23% of the variance in cognition at *r*=0.48, comparable to body and brain markers. Physical activity, diet, alcohol consumption, sun exposure, and local environment emerged as the strongest predictors. A composite brain marker integrating three neuroimaging modalities accounted for 57.7% of the lifestyle–cognition association, while a composite body marker spanning nine physiological systems accounted for 47.8%. Jointly, lifestyle, environment, body, and brain captured 92.6% of age-related cognitive variation. Therefore, integrating lifestyle and environmental factors enables robust prediction of cognition within the UK Biobank cohort, and much of this association is reflected in brain and bodily health. However, the generalizability of these findings to broader populations requires further evaluation.

## 1. INTRODUCTION

Identifying factors that support cognitive health is pivotal for developing strategies to reduce the risk of cognitive impairment. Gerontology research increasingly highlights lifestyle and environmental factors as modifiable targets for maintaining cognitive capacity across adulthood, slowing cognitive decline, and potentially enhancing the effectiveness of pharmacological interventions ^[1–3]^. Healthy diet, regular exercise, adequate sleep, and favourable environmental conditions have all been linked to better cognitive outcomes, potentially through their influence on body and brain health ^[4–10]^. However, most studies examine these factors in isolation and typically report modest associations ^[9,11–13]^. Moreover, they often aim to explain these relationships rather than to develop individual-difference markers that can predict cognitive functioning in new samples outside the model-building process. A cognitive marker that integrates information from diverse lifestyle and environment variables – for example, using machine learning approaches – could provide a more robust characterization of their relationships with cognition and enhance their translational relevance for clinical risk assessment ^[14]^. Determining the domains that contribute most strongly to this marker should also further clarify which aspects of lifestyle and environment hold the greatest promise for supporting cognitive health.

Body health and brain integrity are widely viewed as key pathways through which lifestyle and environmental influences relate to cognition. Behaviors such as smoking, poor nutrition, sleep disturbances, and physical inactivity, as well as adverse environmental exposures like air pollution, have consistently been identified as risk factors for physical illness ^[15,16]^, brain disorders ^[17–19]^, and cognitive decline ^[10,20]^. Growing evidence further suggests that brain structure and function mediate the relationships between physical health, mental health, and cognition ^[21,22]^. In our previous work, we showed that brain characteristics measured across three magnetic resonance imaging (MRI) modalities – diffusion-weighted MRI (dwMRI), resting-state functional MRI (rsMRI), and structural MRI (sMRI) – account for a substantial portion of the shared variance between cognition and body physiology across cardiovascular, pulmonary, renal, hepatic, immune, metabolic, musculoskeletal, and sensory systems ^[23]^.

These neuroimaging modalities index distinct aspects of brain organization. Briefly, dwMRI provides quantitative estimates of white matter microstructure and spatial architecture ^[24]^, rsMRI captures temporal correlations in blood-oxygen-level-dependent (BOLD) signals to characterise functional connectivity across brain regions and networks ^[25]^, and sMRI assesses regional brain anatomy and morphology ^[26]^. All three are widely used in predictive modeling of cognitive performance ^[27–30]^. Accordingly, to understand how well lifestyle and environment variables predict cognition, it is informative to benchmark their predictive performance against neuroimaging measures. More importantly, it remains unclear how brain-based and system-wide physiology-based markers jointly contribute to the lifestyle–environment–cognition relationship. Evaluating their combined influence may provide deeper insight into the biological processes linking lifestyle and environmental exposures to cognitive health.

The physiological and neurobiological processes linking lifestyle, environment, and cognition are dynamic across the lifespan. It is well documented that the accumulation of system-wide deterioration in body and brain health with aging is a major determinant of cognitive functioning and decline. Wear-and-tear processes, including chronic inflammation, insulin resistance, and vascular dysfunction, are associated with cognitive impairment through disruption of the blood–brain barrier, accelerated grey matter atrophy, and the accumulation of white matter lesions ^[31]^. Understanding how diverse lifestyle and environment domains, together with indicators of body and brain health, are linked to age-related variations in cognitive functioning is therefore pivotal for translating predictive markers into clinically meaningful indicators. Quantifying the unique and shared contributions of lifestyle and environmental exposures, system-wide body physiology, and brain structure and function to cognitive aging represents a critical step toward clarifying the extent to which these domains reflect overlapping or distinct pathways.

Here, analysing data from over 10,000 older adults in the UK Biobank, we pursued three aims. First, using machine learning, we quantified how well lifestyle and environmental factors predicted cognitive functioning and identified the domains most strongly associated with this prediction. To contextualize these findings, we compared the predictive performance of lifestyle and environmental factors with that of brain and body phenotypes measured across three neuroimaging modalities – dwMRI, rsMRI, and sMRI – and nine physiological systems, including cardiovascular, pulmonary, renal, hepatic, immune, metabolic, musculoskeletal, hearing, and body and abdominal organ composition. Second, using commonality analyses, we assessed the extent to which body and brain phenotypes accounted for the associations between lifestyle–environment domains and cognitive functioning. Third, recognising age-related variation in cognitive functioning, we conducted an additional commonality analysis to estimate the proportion of age-related differences explained by lifestyle and environmental factors, body and brain markers, and their shared contributions. Figure 1 provides an overview of the research design.

**Figure 1.**
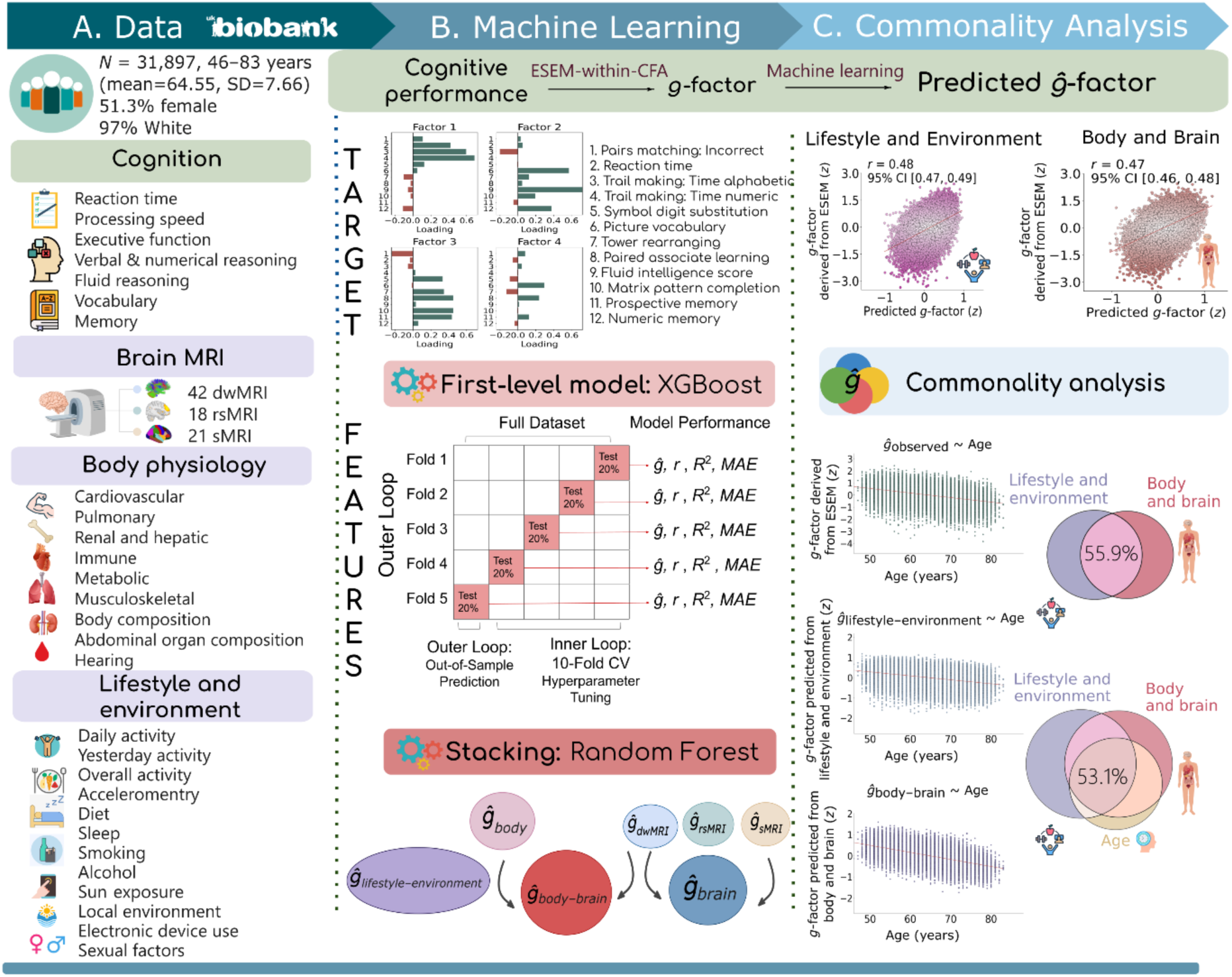
Schematic representation of the research design. **a. Data used in the study.** Cognitive performance was assessed at the first imaging visit (2014– 2019) and comprised twelve scores across eleven cognitive tests. Brain features included 81 phenotypes across three neuroimaging modalities: diffusion-weighted MRI (dwMRI), resting-state functional MRI (rsMRI), and structural MRI (sMRI). Body physiology was represented by nineteen phenotypes spanning nine systems: cardiovascular, pulmonary, renal, hepatic, immune, metabolic, musculoskeletal, hearing, and body/abdominal organ composition. Lifestyle and environment features encompassed twelve domains: activity patterns (daily and yesterday’s physical activity, metabolic equivalent of task (MET), and accelerometry), diet, smoking and alcohol use, sleep, sun exposure, electronic device use, sexual behavior, and local environmental conditions. **b. Two-level machine learning framework.** The target variable was a latent general cognition factor (*g*-factor), estimated from twelve cognitive performance measures using exploratory structural equation modeling within confirmatory factor analysis (ESEM-within-CFA). First-level models (XGBoost) predicted the *g*-factor from each lifestyle/environment domain and each body and brain phenotype. Second-level multimodal stacking integrated predictions across and within modalities: *g*-factor predictions from all first-level models served as input features to Random Forest models that predicted the same target variable. This produced seven composite markers: a lifestyle–environment marker (*ĝ*_lifestyle–environment_), a body marker (*ĝ*_body_), unimodal brain markers from each neuroimaging modality (*ĝ*_dwMRI_, *ĝ*_rsMRI_, *ĝ*_sMRI_), a composite brain marker integrating all brain phenotypes (*ĝ*_brain_), and a combined body–brain marker (*ĝ*_body– brain_). Model performance was assessed using nested cross-validation with five outer folds and ten inner folds. **c. Commonality analysis.** Composite markers and age were included as explanatory variables in linear regression models, with the observed *g*-factor pooled across outer-fold test sets serving as the response variable. To quantify (a) the contribution of body and brain markers to the association between cognition and lifestyle/environment, and (b) the contribution of lifestyle– environment and body–brain markers to age-related cognitive variance, we decomposed the total variance of the observed *g*-factor into components uniquely or jointly explained by each explanatory variable or pair of variables. *dwMRI*, diffusion-weighted MRI; *rsMRI*, resting-state MRI; *sMRI*, structural MRI; *r*, Pearson correlation between observed and predicted *g*-factor values; *R*^2^, coefficient of determination; *MAE*, mean absolute error; *CV*, cross-validation; *CI*, confidence interval. Throughout, “lifestyle and environmental influences” or “characteristics” refer to real-world exposures and behaviors, while “lifestyle and environment domains”, “indices”, or “features” denote their operationalized representations in predictive models.

## 2. RESULTS

### 2.1. Cognition

Cognitive functioning was quantified using a general factor of cognition (*g*-factor) estimated from the covariance among twelve cognitive performance measures ^[32–34]^ via exploratory structural equation modeling within confirmatory factor analysis (ESEM-within-CFA; https://mateuspsi.github.io/esemComp/articles/esem-within-cfa.html). Parallel factor analysis supported a four-factor solution, while the Kaiser–Meyer–Olkin statistic ^[35]^ and Bartlett’s test of sphericity ^[36]^ indicated good factorability. Model goodness-of-fit indices further supported the construct validity of the factor structure ^[37]^. The resulting *g*-factor accounted for 35% of the variance across the twelve cognitive performance measures, consistent with previous findings in the UK Biobank ^[38]^. Supplementary Table S1 lists the cognitive tests and corresponding UK Biobank field numbers, Table S2 reports factor loadings from the ESEM-within-CFA model, and Table S3 summarises model fit statistics.

#### 2.1.1. Predicting cognition from lifestyle and environment

We leveraged a two-level machine learning approach to predict cognition from lifestyle and environment indices spanning twelve domains (Supplementary Table S1) and to derive a composite lifestyle–environment marker. Figure 2A shows the relationship between observed *g*-factor and the composite lifestyle–environment marker, and Figure 2B shows the distribution of predictive performance across the individual lifestyle and environment domains. These domains included daily and yesterday’s physical activity, metabolic equivalent of task (MET), accelerometry, diet, smoking, alcohol consumption, sleep, sun exposure, electronic device use, sexual behavior, and local environmental characteristics. Integrating all domains into a composite lifestyle–environment marker led to predictive performance at *r*_mean_ = 0.479 (95% confidence interval, CI [0.47, 0.487]), accounting for 23% of the variance in cognition and outperforming individual lifestyle and environment domains (Figure 2A; see Supplementary Tables S4–S6 for full statistics).

**Figure 2.**
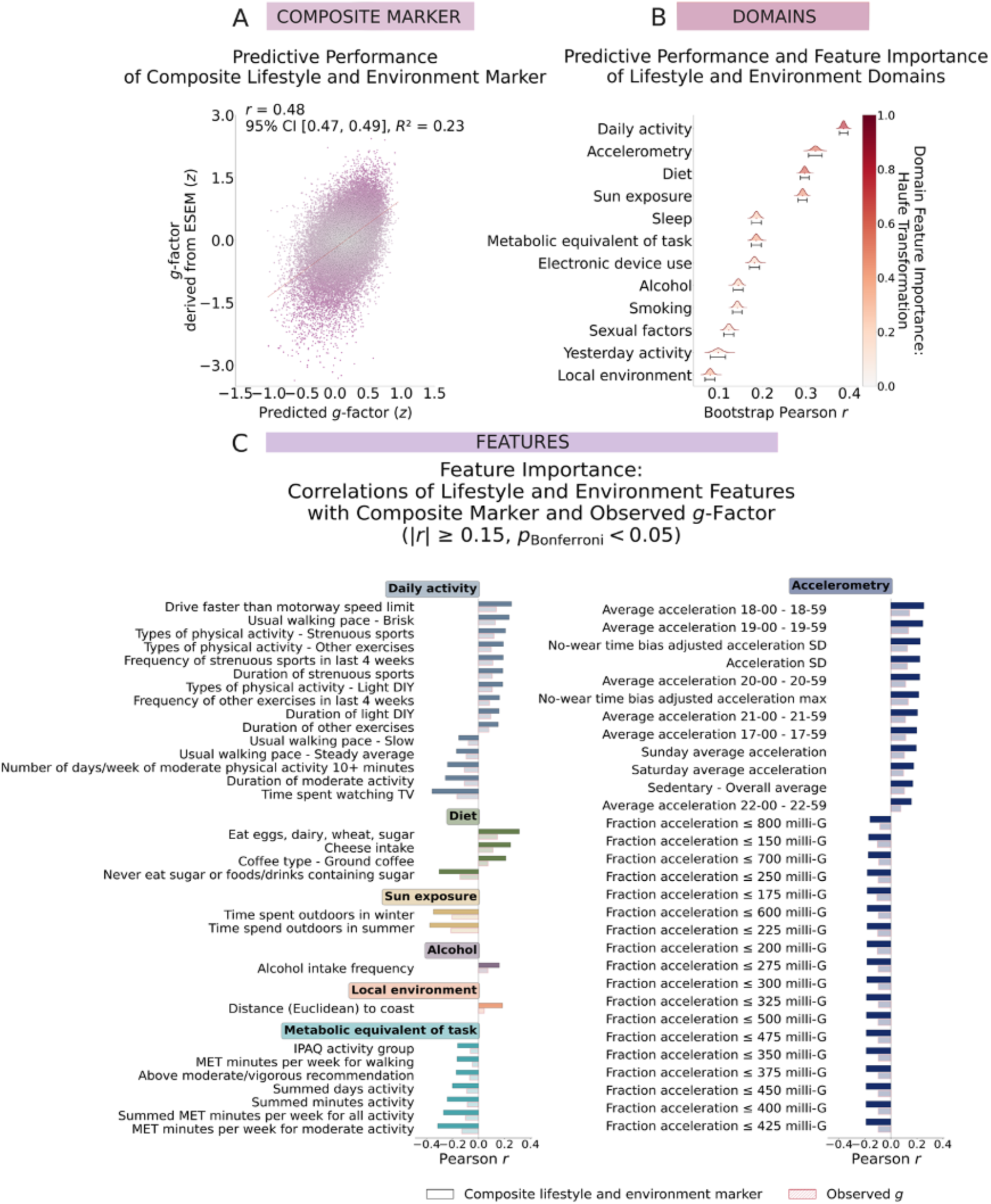
Predictive performance and feature importance of lifestyle and environment domains. **a.** Scatterplot showing the relationship between the observed *g*-factor (derived via ESEM-within-CFA) and the *g*-factor predicted by the stacked model integrating all twelve lifestyle and environment domains (the composite lifestyle–environment marker). Pearson *r* represents the mean correlation between observed and predicted *g*-factor values across the five outer-fold test sets, with 95% confidence intervals (CIs) estimated via bootstrap resampling of the pooled data. **b.** Ridgeline plots depicting the bootstrap distribution of predictive performance for each lifestyle and environment domain, along with 95% CIs. The color fill for each domain reflects its association with the composite lifestyle–environment marker, quantified as the Pearson correlation between the *g*-factor predicted by the individual domain and the *g*-factor predicted by the stacked model integrating all domains (Haufe transformation). **c.** Bar plots indicating the direction and magnitude of Pearson correlations between individual lifestyle and environment features and (1) the *g-*factor predicted by the stacked model (darker bars) and (2) the observed *g-*factor (lighter hatched bars). Only correlations that remained significant after Bonferroni correction (*p* < 0.05) and with absolute values |*r*| ≥ 0.15 are shown. A full list of feature importance values is provided in Supplementary Table S7.

Individually, lifestyle and environment domains explained between 0.6% and 15% of the variance in cognition, with mean Pearson correlations (*r*_mean_) between observed and predicted *g*-factor scores ranging from 0.08 to 0.39 (Figure 2B; Supplementary Tables S4, S5). Daily activity yielded the strongest predictive performance (*r*_mean_ = 0.39, 95% CI [0.377, 0.396]), followed by accelerometry-based measures (*r*_mean_ = 0.32, 95% CI [0.307, 0.337]). In contrast, local environmental characteristics and previous-day activity showed the weakest associations, with *r*_mean_ = 0.10 (95% CI [0.08, 0.115]) and 0.08 (95% CI [0.068, 0.091]), respectively (Figure 2B).

To identify the domains and features most relevant to cognition, we conducted a feature importance analysis using the Haufe Transformation ^[39]^. Figure 2C shows the direction and magnitude of associations between individual lifestyle and environment features and both the predicted composite lifestyle–environment marker and the observed *g*-factor (Supplementary Table S7). Domain-level importance was quantified as the Pearson correlation between the *g*-factor predicted from each domain and the *g*-factor predicted by the stacked model (i.e., the composite lifestyle–environment marker). Within individual domains, feature-level importance was assessed by correlating each feature with the composite lifestyle–environment marker (*r*_composite_). To verify the directionality of the Haufe-derived feature importance estimates, we additionally computed zero-order correlations between each feature and the observed *g*-factor.

At the domain level, daily activity (*r*_composite_ = 0.78, *p* < 0.0001), diet (*r*_composite_ = 0.63, *p* < 0.0001), physical activity measured by accelerometry (*r*_composite_ = 0.57, *p*<0.0001) and sun exposure (*r*_composite_ = 0.47, *p* < 0.0001) showed the strongest associations with the composite marker, whereas previous-day activity (*r*_composite_ = 0.14, *p* < 0.0001) and sexual behavior (*r*_composite_ = 0.17, *p* < 0.0001) exhibited the weakest associations. At the feature level, the strongest associations with the composite lifestyle–environment marker were observed for activity-related measures (daily activity, MET, accelerometry) as well as dietary factors, alcohol consumption, sun exposure, and local environment (Figure 2C).

Positive associations (*r*_composite_ = 0.15 to 0.25, *p* < 0.0001) included faster self-reported driving and walking pace, greater frequency of strenuous sports and light do-it-yourself (DIY) activities, and higher accelerometry-derived activity during evenings and weekends. In contrast, negative associations (*r*_composite_ = –0.35 to –0.15, *p* < 0.0001) were demonstrated for television viewing time, self-reported moderate activity level, slower walking pace, and a greater fraction of time spent below 800 milli-gravities acceleration. All MET indices, including weekly durations of vigorous and moderate activity, walking, and overall daily activity, were negatively correlated with the composite marker (*r*_composite_ < –0.31, *p* < 0.0001) (Figure 2C). Notably, the directions of Haufe-derived feature importance estimates were consistent with those obtained using zero-order correlations between each feature and the observed *g*-factor.

Among dietary factors, intake of eggs, dairy, wheat, and sugar (*r*_composite_ = 0.31, *p* < 0.0001), cheese (*r*_composite_ = 0.25, *p* < 0.0001), and ground coffee (*r*_composite_ = 0.21, *p* < 0.0001) showed positive associations with the composite marker of cognition, as did alcohol intake frequency (*r*_composite_ = 0.16, *p* < 0.0001). In contrast, reporting never consuming sugar or sugar-containing foods and drinks was negatively associated with the composite marker (*r*_composite_ = – 0.30, *p* < 0.0001). For environmental characteristics, longer time spent outdoors during both winter (*r*_composite_ = – 0.35, *p* < 0.0001) and summer (*r*_composite_ = –0.37, *p* < 0.0001) was associated with lower cognition scores, whereas greater distance from the coast was associated with higher scores (*r*_composite_ = 0.18, *p* < 0.0001) (Figure 2C).

#### 2.1.2. Predicting cognition from body and brain phenotypes

To benchmark the predictive performance of the lifestyle–environment marker, we compared its performance against composite body and brain markers using bootstrapping. The composite body marker was derived by first training 19 base models, each predicting the *g*-factor from a single body phenotype across nine systems (cardiovascular, pulmonary, renal, hepatic, immune, metabolic, musculoskeletal, hearing, and body/abdominal composition) and then integrating these models via multimodal stacking. Similarly, unimodal and composite brain markers were obtained by training base models on each of the 81 brain phenotypes from dwMRI, rsMRI, and sMRI modalities and combining them within and across modalities. Finally, a combined body–brain marker was constructed by integrating all body and brain phenotypes.

The composite lifestyle–environment marker performed similarly to the composite brain marker and the combined body–brain marker but significantly outperformed the composite body marker and all unimodal brain markers, as well as individual body and brain phenotypes. Figure 3A shows the bootstrap distributions of differences in predictive performance between the lifestyle–environment marker and the body and brain markers, and Figure 3B illustrates the relationships between observed and predicted *g*-factor scores for the composite markers. Supplementary Table S6 reports bootstrap differences in performance metrics, and the cross-validated and bootstrapped performance metrics for body and brain phenotypes and composite markers are provided in Supplementary Tables S8 and S9, respectively.

**Figure 3.**
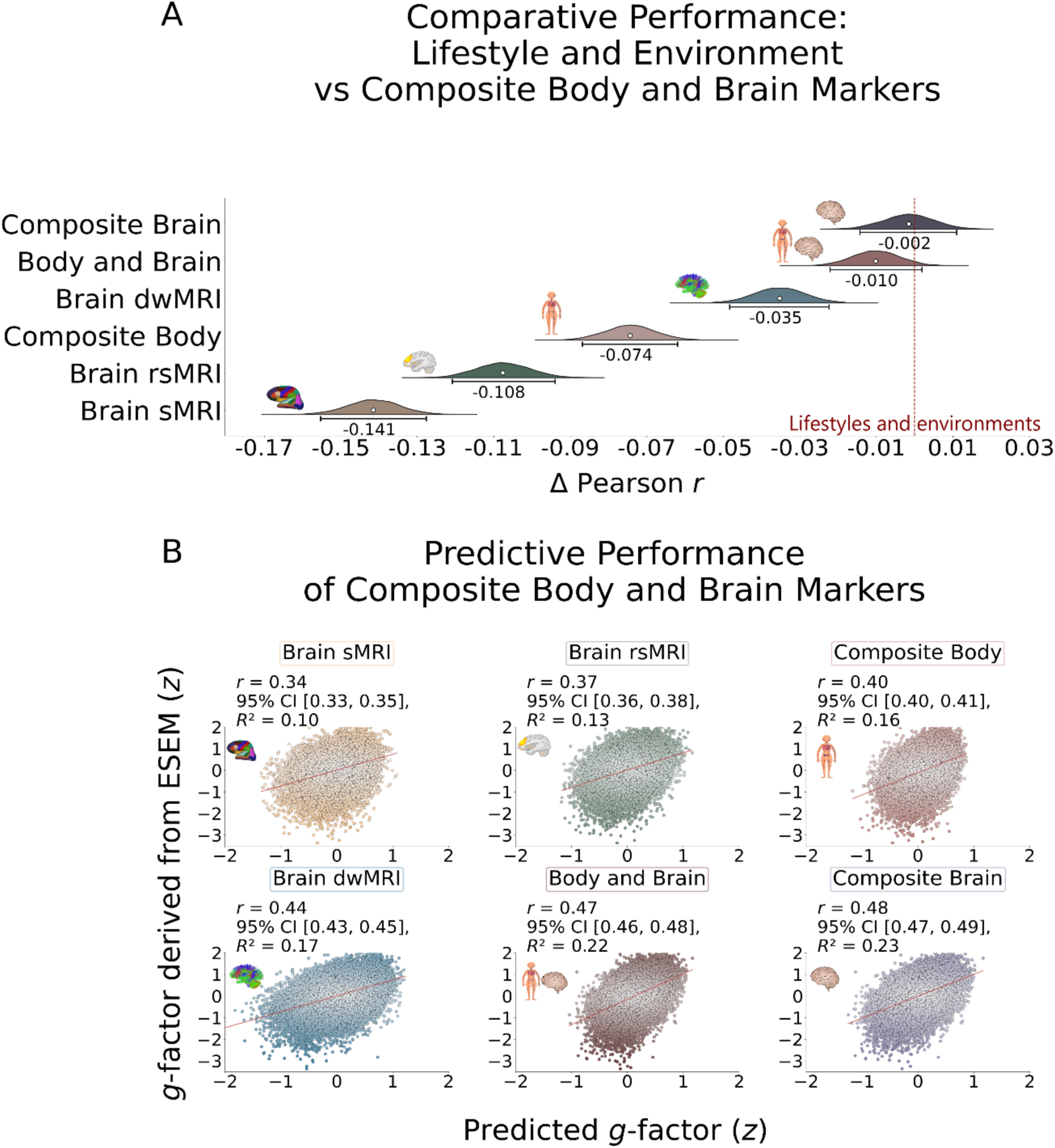
Absolute and comparative predictive performance of body and brain phenotypes. **a.** Kernel density estimation plot showing the bootstrap distribution of differences (*Δ*) in Pearson *r* between the composite lifestyle–environment marker and each of the following: the composite body marker, unimodal brain markers, the composite brain marker, and the combined body–brain marker, together with their 95% confidence intervals (CIs). **b.** Scatterplots depicting the relationship between the observed *g*-factor (derived via ESEM-within-CFA) and the *g*-factor predicted by stacked models generating four composite markers: the composite body marker, the unimodal brain markers, the composite brain marker, and the combined body–brain marker. Pearson *r* represents the mean correlation between observed and predicted *g*-factor values across the five outer-fold test sets, with 95% CIs estimated via bootstrap resampling of the pooled data.

### 2.2. Commonality Analyses

We employed commonality analysis to (a) determine the proportion of the lifestyle– environment–cognition relationship accounted for by body and brain markers, and (b) quantify the proportion of age-related cognitive variance captured by the composite lifestyle– environment and body–brain markers. The observed *g*-factor served as the response variable, with *g*-factors predicted from lifestyle/environment, body, brain, or age as explanatory variables. Figure 4 summarizes the unique and shared contributions of the lifestyle– environment, body and brain markers, and age to cognitive variance, with detailed results provided in Supplementary Tables S10 and S11.

**Figure 4.**
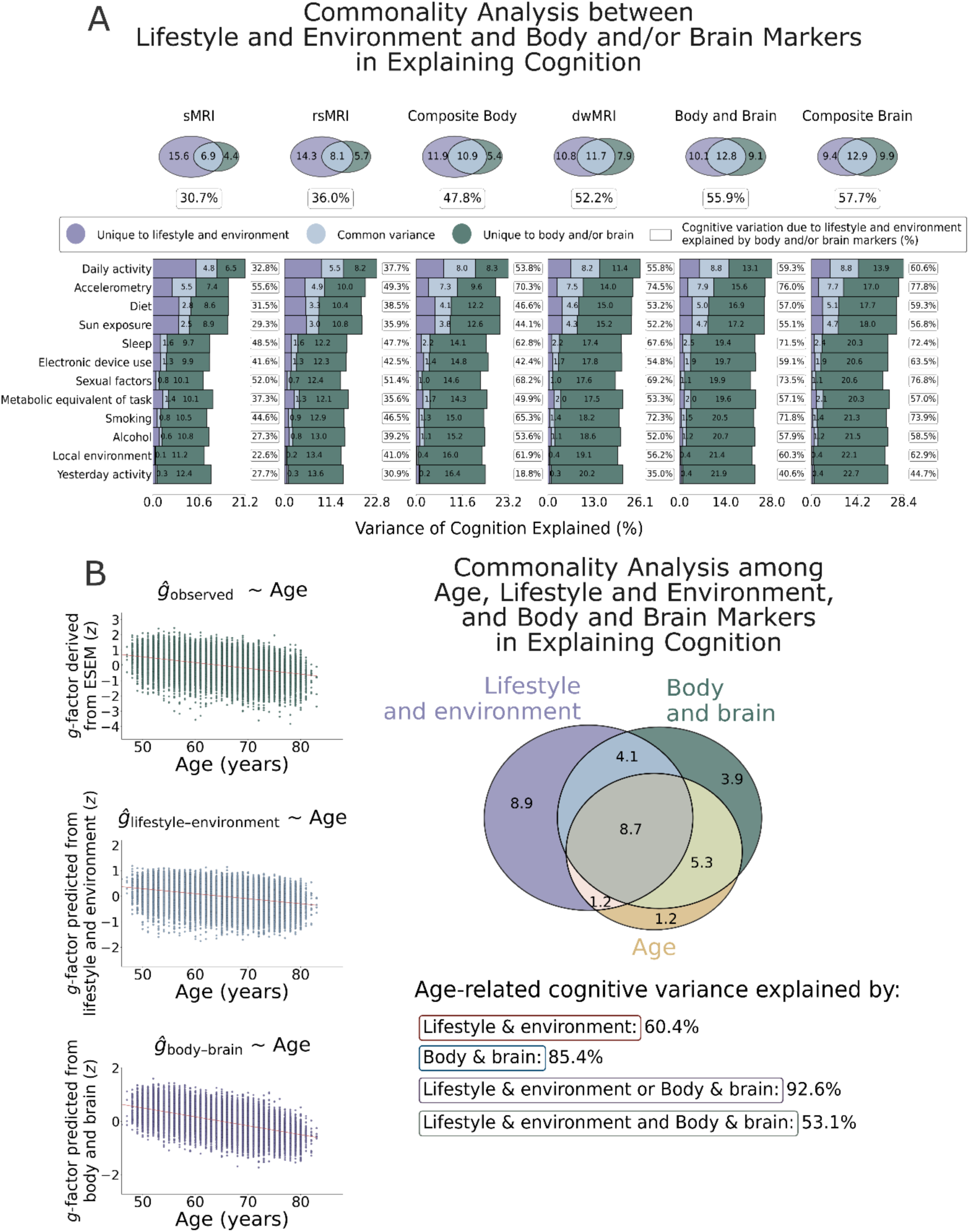
Commonality analyses. **a.** Commonality analysis between the lifestyle–environment and body and brain markers in explaining cognition. Venn diagrams (top) depict the unique and shared contributions of the composite lifestyle–environment marker and individual body or brain markers to cognitive variance. Stacked bar plots (bottom) show the unique and shared contributions of each lifestyle/environment domain and a given body/brain marker to cognitive variance. Percentages in each box indicate the proportion of lifestyle–environment–cognition covariation explained by a given body or brain marker, calculated as: 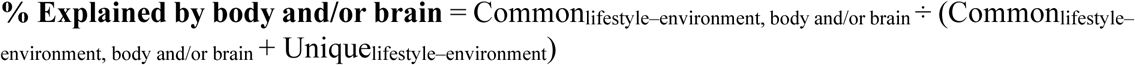 Unique variance refers to the proportion (%) of variance in the *g*-factor explained exclusively by one variable; common variance reflects the proportion (%) explained jointly by two variables **b.** Commonality analysis among age, the composite lifestyle–environment marker, and the composite body–brain marker in explaining cognition. A Venn diagram illustrates the unique and shared contributions of age, the lifestyle–environment marker, and the body–brain marker to cognitive variance. Unique variance refers to the proportion (%) of variance in the *g*-factor explained exclusively by one explanatory variable; common variance reflects the proportion (%) explained jointly by two or more explanatory variables. Percentages of age-related cognitive variance explained were calculated as: 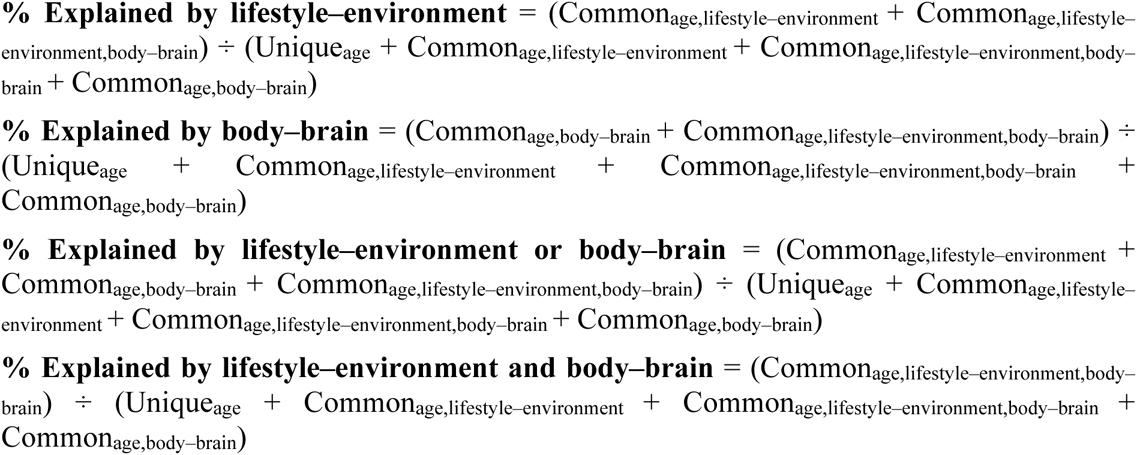 Scatterplots show the observed *g*-factor (derived via ESEM-within-CFA) and predicted *g*-factor values from the stacked lifestyle–environment and body–brain models, plotted as a function of age.

#### 2.2.1. Commonality analysis between the lifestyle–environment and body and brain markers in explaining cognition

To quantify the proportion of the lifestyle–environment–cognition association captured by body and brain markers, we calculated the ratio of shared variance. Specifically, we divided the variance in the observed *g*-factor jointly explained by the lifestyle–environment marker and a given body or brain marker by the total variance explained by the lifestyle–environment marker alone.

Most composite markers accounted for roughly half of the relationship between lifestyle/environment and cognition. The composite brain marker explained the largest proportion at 57.7%, followed closely by the composite body–brain marker at 55.9%. The composite body marker accounted for 47.8%. Among the three neuroimaging modalities, dwMRI captured the largest share of lifestyle/environment-related cognitive variance (52.2%), followed by rsMRI (36%) and sMRI (30.7%) (Figure 4A).

Overall, the composite brain and body–brain markers explained over 75% of the relationship between cognition and accelerometry, and approximately 60% of its association with daily activity. For several other lifestyle domains, specifically sexual factors, sleep, and smoking, the composite body, brain, body–brain, and dwMRI markers accounted for a substantial portion (68–77%) of the association with cognition. The explanatory power of sMRI and rsMRI markers was moderate but meaningful, ranging from 27.7% to 55.6% and from 30.9% to 51.5%, respectively, across all lifestyle and environment domains (Figure 4A; Supplementary Table S10).

#### 2.2.2. Commonality analysis among age, the composite lifestyle–environment marker, and the composite body–brain marker in explaining cognition

We quantified the contributions of the lifestyle–environment and body–brain markers to age-related cognitive variance as percentage ratios, where the numerator represented the variance in the observed *g*-factor jointly explained by age and a given marker, and the denominator corresponded to the total variance explained by age.

Individually, the composite lifestyle–environment and body–brain markers accounted for 60.4% and 85.4% of the age-related variance in cognition, respectively. When considered together, they explained 92.6% of this variance, with a 53.1% overlap between the two markers (Figure 4B; Supplementary Table S11).

## 3. DISCUSSION

In this study, we evaluated the predictive utility of multiple lifestyle and environment domains for cognitive functioning and examined their associations with cognitive health and aging. By integrating information across twelve domains in a large cohort of older adults, we derived a composite lifestyle–environment marker of cognition (*r*_lifestyle–environment_ = 0.48; *r*_composite body physiology_ = 0.40, *r*_composite brain_= 0.48), highlighting the contribution of lifestyle and environmental factors to cognitive performance alongside other biological and physiological measures within the UK Biobank sample. We further showed that approximately half (55.9%) of the variance in cognition explained by lifestyle and environmental factors is captured by indicators of bodily health and brain structure and function. Finally, we demonstrated that jointly considering lifestyle and environment alongside body and brain measures explains 92.6% of age-related variance in cognition, with a substantial shared component (53.1%).

### 3.1. Lifestyle and Environment Domains Rival Body and Brain Phenotypes in Predicting Cognition

Lifestyle and environmental exposures are well-established predictors of cognitive outcomes in older adults ^[10,20,40,41]^. Rather than examining these factors individually, we used machine learning to derive an integrated cognitive marker that combined information across many lifestyle and environment variables. This marker showed out-of-sample predictive performance comparable to composite brain and combined body–brain markers, and outperformed markers based on single neuroimaging modalities or system-wide physiology. Although individual features had modest effects, multimodal stacking produced a robust marker explaining 27% of cognitive variance, consistent with findings from the Cognitive Function and Ageing Study – Wales ^[42]^. Integrating diverse exposures in this way can help address key challenges in lifestyle and environmental research and offers a scalable framework for incorporating behavioral and environmental data into cognitive prediction models.

Our findings further extend previous work by identifying the lifestyle and environment domains most strongly associated with cognition and evaluating their comparative predictive performance. Among the twelve domains examined, daily activity captured through self-report emerged as the strongest predictor of cognitive functioning, followed by diet, accelerometry-derived physical activity, sun exposure, overall activity assessed with MET, and local environmental characteristics. Features related to alcohol consumption also showed substantial associations with cognition. These domains may therefore represent promising targets for supporting cognitive health in later life and provide a foundation for strategies aimed at maintaining both physical and brain health through lifestyle modification. Evaluating the impact of such approaches will be essential to determine whether modifying these factors leads to measurable improvements in cognitive outcomes. It is important to note, however, that our machine learning framework was designed for out-of-sample prediction rather than causal inference. As such, it does not establish the directionality of the associations between cognition and each lifestyle or environmental feature. Our interpretation, therefore, assumes that these relationships may reflect influences in either direction or mutually reinforcing processes.

The three activity domains, including daily activity, accelerometry, and MET, are among the top domains contributing to the prediction of cognition. We demonstrate the utility of accelerometry as an objective measure of activity level, distinct from daily-activity questionnaires. Future research may benefit from incorporating a greater proportion of instrument-based measures, which could reduce potential bias associated with self-reported behavioral assessments. At the feature level, positive associations (*r* > 0.15) for daily physical activity were observed for faster driving speed, faster self-reported walking pace, and engagement in strenuous sports and light DIY activities. In contrast, a slower walking pace, greater television viewing time, and self-reported moderate activity level were associated with poorer cognitive performance. For accelerometry-derived measures, activity during evenings and weekends showed positive associations with cognition, whereas a higher proportion of time spent at acceleration magnitudes ≤800 milli-gravities showed the opposite pattern. All MET indices were linked to lower cognitive scores.

Driving behavior may serve as an ecologically valid indicator of cognitive health, as safe and efficient driving relies on intact visuospatial processing, executive function, attention, and memory ^[43–45]^. Reductions in driving frequency, route complexity, and spatial range have been linked to mild cognitive impairment ^[46,47]^; thus, faster driving speed may reflect preserved cognitive and sensorimotor function. Walking speed shows a similarly robust relationship with cognition. Reduced walking speed often precedes cognitive decline and predicts incident dementia, suggesting it may represent an early behavioral manifestation of Alzheimer’s pathology ^[45,48–51]^, with longitudinal studies demonstrating parallel trajectories of gait velocity and cognitive performance ^[52]^. This aligns with the Motoric Cognitive Risk Syndrome framework, which conceptualizes co-occurring subjective cognitive complaints and motor signs such as slow gait as a pre-dementia state ^[53]^. These associations are thought to share common neuropathological substrates, including reduced frontal white matter integrity, lower temporal and total cortical grey matter volumes, smaller hippocampal volumes, and enlarged ventricles ^[54–56]^.

Engagement in strenuous sports and higher overall physical activity have consistently been linked to better cognitive outcomes and reduced dementia risk ^[57,58]^. Prior work suggests that accelerations above 425 milli-gravities correspond to vigorous physical activity ^[59,60]^; therefore, the negative correlation between time spent at ≤800 milli-gravities and cognition aligns with evidence that reduced engagement in strenuous activity is associated with poorer cognitive performance. These associations are likely bidirectional. On one hand, higher activity levels may reflect preserved cognitive function, motivation, and overall bodily health – an important determinant of brain integrity and a well-established protective factor against cognitive impairment ^[61]^. On the other hand, physical activity may support brain health and neuroplasticity through increased production of neurotrophic factors such as brain-derived neurotrophic factor and insulin-like growth factor-1 ^[62,63]^, exercise-induced myokines fibronectin type III domain-containing protein 5 (FNDC5) and Irisin ^[64,65]^, alongside enhanced neurogenesis, gliogenesis, synaptogenesis, angiogenesis, and modulation of neurotransmission ^[6,66–68]^. Light DIY activities and greater activity during evenings and weekends may additionally capture aspects of social engagement, autonomy, and purposeful behavior, all of which have been associated with cognitive resilience, wellbeing, and reduced stress ^[69–71]^. Supporting this view, studies show that social contacts and lower depressive symptoms mediate the relationship between accelerometer-derived physical activity and cognitive health ^[72]^. Such activities may therefore index not only physical exertion but also cognitive reserve, social participation, and structured daily routines ^[73–76]^. Critically, these findings align with theories proposing that intellectually and physically challenging, socially engaging activities are associated with better cognition across multiple domains, including executive function as well as crystallized and fluid intelligence ^[77,78]^.

By contrast, greater television viewing time was associated with poorer cognition, consistent with prior findings in the UK Biobank ^[9]^. Interestingly, accelerometer-derived sedentary time showed the opposite pattern, and self-reported computer-based screen use exhibited a weak but positive association with cognition (*r* = 0.069, *p* < 0.0001), highlighting that sedentary behaviors are heterogeneous rather than uniformly detrimental. Television viewing typically involves cognitively passive engagement with high sensory input and minimal executive demand ^[79]^. Prior work links excessive television viewing to neurobiological alterations, including reduced axonal and dendritic density in regions supporting language, communication, and memory ^[79,80]^, as well as reduced total grey matter and lower grey matter volume in frontal and entorhinal cortices ^[81]^. Conversely, computer use is more cognitively active, often involving information processing, problem-solving, and digital literacy, and may reflect higher educational engagement and the use of digital tools as sources of information ^[82]^. Overall, digital engagement has been associated with better executive functioning, reasoning, and protection against cognitive impairment in older adults ^[83]^, with particular benefits for visual–motor skills and task-switching ^[82,84]^.

Importantly, self-reported physical activity measures are known to be susceptible to reporting bias. Indeed, questionnaire- and accelerometer-derived activity measures show only weak to moderate correspondence, with stronger agreement among individuals of higher socioeconomic status ^[85–88]^. Cognition is another key factor associated with difficulties in recalling past activities and with over- or underreporting of daily physical activity, especially among older individuals ^[89]^. However, the observation that other self-reported activity measures in the present study showed positive associations with cognition suggests that reporting bias alone is unlikely to fully account for the negative association observed for MET-derived indices. This discrepancy may instead reflect differences in how activity constructs are operationalized (e.g., intensity-weighted MET estimates versus frequency- or duration-based measures). Consistent with prior findings in the UK Biobank ^[11]^, these results highlight that both measurement characteristics and sample composition can influence observed associations. In addition, associations between physical activity and cognition may also be shaped by collider bias, because both lifestyle factors and cognitive performance are related to the likelihood of volunteering in the UK Biobank ^[90]^.

Other lifestyle and environmental factors most strongly related to cognition came from diet, alcohol consumption, sun exposure, and local environmental characteristics. For dietary factors, low to moderate positive correlations were observed for the consumption of eggs, dairy, including cheese, wheat, sugar, and ground coffee. These associations should be interpreted cautiously, as they may partly reflect the structure of the questionnaire. While cheese and coffee consumption were assessed as separate questionnaire items and showed positive associations with cognition, both the positive association with eating eggs, dairy, wheat, and sugar and the negative associations with reporting never eating wheat products, dairy products, eggs or foods containing eggs, and sugar or foods/drinks containing sugar were derived from the same question: *“Which of the following do you never eat?”*. Thus, the positive association reflects not reporting avoidance of these foods *(“I eat all of the above”*), whereas the negative associations indicate that participants who explicitly reported avoiding individual foods tended to have lower cognition scores. These findings may not indicate that consumption of these foods is detrimental to cognition. Instead, it is possible that individuals who avoid multiple foods may differ from other participants in underlying health conditions, medically advised dietary restrictions, or broader dietary and lifestyle characteristics, which may also be associated with cognition. Although biological, behavioral, and broader dietary pattern mechanisms remain possible, the structure of the questionnaire, particularly the absence of information on quantity and frequency of consumption and the categorical nature of the responses, limits our ability to distinguish among these potential explanations. Accordingly, these findings are best interpreted as associations with reported dietary behavior rather than evidence for effects of individual foods on cognition.

The potential cognitive benefits of cheese may relate to its content of fermentation-derived bioactive peptides rich in tryptophan and cysteine, lactic acid bacteria, vitamin B12, calcium, and other nutrients ^[91,92]^. Previous findings on cheese intake and cognition in older adults have been mixed, with some studies reporting inverse associations with cognitive impairment ^[91,93–96]^, while others have reported non-significant associations or nonlinear relationships with cognitive decline and dementia risk ^[96–98]^. A UK Biobank study showed that daily cheese intake predicted better fluid intelligence scores over time ^[99]^, and another longitudinal study found positive associations with executive function and verbal fluency ^[100]^. Importantly, results vary across regions and depend on both the amount and type of cheese consumed ^[98,101–103]^.

Numerous studies have reported associations between coffee consumption and cognitive performance ^[104,105]^ as well as dementia risk ^[106–109]^, with the lowest risk observed in hypertensive individuals and among those who consume ground coffee ^[106]^. The potential beneficial effects of ground coffee may relate to its higher content of caffeine and other biologically active compounds ^[110,111]^, including polyphenols such as chlorogenic acid, which exert protective effects on neurons, microglia, and cognition ^[112,113]^. Caffeine is thought to be the primary component responsible for these positive effects, as its neuroprotective properties are well established ^[114–116]^. In contrast, instant coffee tends to contain lower levels of active compounds and higher levels of toxins such as acrylamide and heavy metals ^[117,118]^, and it is often consumed as part of commercial beverages rich in processed ingredients, including sugars, powdered cream, and other additives, that may offset the beneficial effects of bioactive compounds ^[110,119,120]^. Notably, we also observed a negative correlation between instant coffee consumption and cognition, although the effect size was small (*r* = 0.13, *p* < 0.0001). Instant coffee intake has previously been linked to chronic obstructive pulmonary disease ^[120]^ and metabolic syndrome ^[119]^, both of which are associated with poorer cognition ^[23]^, and has causal negative effects on telomere length ^[121]^.

Previous research on alcohol consumption and cognition is contentious. Current public health guidance emphasises that there is no safe dose of alcohol ^[122,123]^, and the detrimental effects of even small amounts on cognition and brain health are broadly acknowledged ^[124–127]^. Although several studies ^[128–131]^, including analyses from the UK Biobank ^[9,132,133]^, have identified nonlinear J- or U-shaped associations between alcohol use and cognitive decline ^[133–136]^, as well as linear positive relationships between light-to-moderate alcohol consumption and cognitive functioning ^[42]^, these patterns tend to attenuate when controlled for income, education, intelligence, and cultural factors ^[126,137]^, and are thought to reflect artefacts of selection and reporting bias ^[133]^. Such associations are therefore often interpreted as proxies for socioeconomic status ^[138–140]^ and social engagement ^[141–143]^, both of which are independently related to cognitive functioning in later life ^[144–147]^.

For sun exposure, greater time spent outdoors during both winter and summer was associated with poorer cognitive performance. Evidence linking sunlight exposure with cognition is equivocal. While some studies using linear models report positive associations between long-term sunlight exposure and domains such as new learning, visual memory, and sustained attention, alongside poorer reaction time ^[148]^, others describe J-shaped relationships with cognitive decline and brain atrophy, with the direction of association changing at approximately 1.5–2 hours of exposure ^[149–151]^. Proposed biological mechanisms include increases in blood flow and body temperature, as well as direct tissue heating via light penetration, which may ultimately raise brain temperature ^[150,152–154]^. Although ultraviolet radiation has also been suggested to exert protective effects against dementia ^[155]^, excessive heat and UV exposure can influence brain morphology, electrophysiology, and blood–brain barrier integrity through inflammatory responses, neuroimmune and transcriptomic alterations, oxidative stress, and changes in neuronal metabolism and neurotransmission ^[156–164]^. At the same time, higher levels of outdoor activity are consistently linked to lower dementia risk, better cognitive performance, mental attention restoration, improved cerebral blood flow, larger hippocampal and total grey matter volumes, and fewer white matter hyperintensities ^[165–167]^. Taken together, these findings suggest that associations between sunlight exposure and cognition may depend less on sunlight itself and more on the behavioral factors accompanying outdoor time, including the types of activities performed, occupational demands, or broader lifestyle characteristics. Notably, the UK Biobank measure used in the present study captures overall time spent outdoors but does not distinguish between recreational and occupational exposure or characterize the surrounding environment, such as natural versus urban settings. Thus, the observed negative association may reflect a combination of environmental, occupational, socioeconomic, health, or lifestyle factors that could not be disentangled in the present analysis. In addition, we did not empirically test or stratify these associations by education or occupation and therefore cannot determine whether occupational exposure contributes to the observed pattern. Moreover, the two studies reporting negative associations between prolonged sunlight exposure and cognition ^[149,150]^ were conducted in the UK Biobank cohort, which primarily includes white UK residents, indicating that cohort-specific patterns may contribute to these findings. Accordingly, reported time spent outdoors should be interpreted as a proxy for a complex set of environmental and behavioral exposures rather than as evidence of a direct adverse effect of sunlight on cognition.

A similarly complex pattern emerged for local environmental characteristics. Greater distance from the coast showed a positive association with cognition. Although proximity to coastal environments is often linked to better mental health, general health, and cognitive outcomes ^[168–170]^, evidence within the UK Biobank suggests more nuanced associations, indicating that environmental benefits may depend on contextual factors such as urbanicity, rurality, deprivation, socioeconomic conditions, or remoteness from major urban centres and access to resources. One study reported that greater coastal distance was associated with better abstract reasoning but poorer executive function ^[147]^, while another found that coastal distance and deprivation partially mediated the relationship between education and cognition, with education and socioeconomic factors explaining substantially more variation overall ^[171]^. Hence, in the UK context, distance from the coast may act as a proxy for sociodemographic characteristics rather than reflecting direct environmental effects on cognitive functioning.

The UK Biobank cohort is well known to differ systematically from the general population: participants tend to be healthier, more educated, and more socioeconomically advantaged, and they engage less frequently in behaviors linked to adverse health outcomes ^[172–176]^. For instance, individuals who participated in neuroimaging assessments show a pronounced “healthy volunteer” profile across cognitive, physiological, neurological, and mental health measures compared with those who did not undergo scanning ^[173]^. As a result, effect sizes may be attenuated, and in some cases, the direction of associations may not fully reflect patterns in the general population ^[173,174]^, with socio-behavioral variables being particularly vulnerable to such participation bias ^[175]^. Collectively, these limitations warrant caution when interpreting and generalising the results of the study.

### 3.2. Body and Brain Markers Capture Nearly Half of the Lifestyle–Environment– Cognition Relationship

Leveraging multimodal data spanning lifestyle and environmental factors, system-wide physiology, and brain neuroimaging, we constructed composite markers to quantify the extent to which the associations between cognitive functioning and body and brain systems overlap with lifestyle and environmental exposures. Both body and brain markers emerged as major contributors to the lifestyle–environment–cognition relationship, with the greatest explanatory power observed when information was integrated across neuroimaging modalities (57.7%) and when neuroimaging was combined with system-wide physiology (55.9%). Notably, incorporating body physiology alongside brain phenotypes did not substantially increase the contribution beyond that achieved by neuroimaging alone, consistent with our previous finding that brain measures capture a large portion of cognition-related variance associated with bodily health ^[23]^.

Across individual modalities, system-wide physiology and neuroimaging captured between 30.7% and 52.2% of the lifestyle–environment–cognition relationship, with the largest overlaps observed for dwMRI, followed by rsMRI and sMRI. These neuroimaging modalities are widely used in predictive modeling of cognitive performance ^[27–30,177]^, and our findings provide quantitative support for evidence that lifestyle and environmental factors relate to cognition through biological processes reflected in brain structure and function, either directly or indirectly via bodily physiology ^[16–18,178,179]^. Importantly, domain-specific analyses showed that body and brain markers captured over 60% of the association between cognition and physical activity, reinforcing prior evidence that physical activity exerts broad effects on both systemic physiology and brain health ^[180,181]^. Substantial explanatory contributions were also observed across other lifestyle domains, such as sleep, smoking, and sexual factors, indicating that body and brain systems account for a large proportion of covariance between diverse behavioral exposures and cognitive functioning. Proposed mechanisms include but are not limited to pathways involving inflammation, neurotransmission, neurotrophic signalling, neurogenesis, hypoxia, antioxidant defence, immune and vascular function, stress response, and neuronal energy metabolism ^[182–190]^.

Consistent with evidence that the brain represents a central pathway linking physical health with cognitive functioning ^[23]^, our results suggest that associations between lifestyle and environmental exposures and cognition are embedded within neurobiological processes that are closely intertwined with systemic physiology. We also extend prior work demonstrating that body systems and brain structure account for lifestyle–mental health associations ^[21]^ by providing quantitative evidence that these biological domains also underpin the relationship between lifestyle and cognitive functioning.

Collectively, these findings support the utility of neuroimaging and system-wide physiological indices for constructing predictive markers that capture key biological pathways linking external exposures with cognitive outcomes. Future research is required to establish causal relationships, clarify how lifestyle factors relate to changes in body and brain health, and delineate the mediating roles of these systems in shaping cognitive trajectories. Elucidating cellular and molecular mechanisms underlying these associations will be essential for advancing beyond systems-level markers derived from neuroimaging and physiology.

### 3.3. Lifestyle–Environment and Body–Brain Markers Overlap with a Substantial Proportion of Age-Related Cognitive Variation

This study is the first to quantify the relative contributions of lifestyle and environmental factors alongside body and brain characteristics to cognitive aging using integrated composite markers. When considered together, the composite lifestyle–environmental and body–brain markers overlapped considerably with age-related cognitive variation, accounting for 92.6% of the variance associated with chronological age, with a substantial shared component (53.1%).

Importantly, this estimate should not be interpreted as evidence that these markers explain cognition beyond chronological age or as evidence of a causal contribution to age-related cognitive decline. Rather, it indicates that these lifestyle–environment and body–brain markers capture variance in cognitive functioning that substantially overlaps with chronological age. Because many body and brain measures are themselves sensitive to chronological age, this overlap may partly reflect their role as biological correlates of aging. Each domain also showed substantial overlap with age-related cognitive variation when examined individually (60.4% for lifestyle and environment; 85.4% for body and brain), highlighting the interconnected relationships between behavioral and biological characteristics and cognitive aging.

Myriad studies demonstrate that lifestyle choices influence trajectories of cognitive aging and support lifestyle modification as a potential avenue for delaying cognitive impairment ^[42,187,191–193]^. Our findings extend this perspective by quantifying the combined contribution of behavioral and biological domains and showing that their overlap is considerable. Critically, this convergence suggests that lifestyle and environmental exposures may operate partly through body and brain pathways while also contributing independent variance, reinforcing the importance of considering these domains jointly rather than in isolation. Overall, these results emphasize that cognitive aging reflects the cumulative influence of behavioral, environmental, and biological factors. Integrative approaches that address both lifestyle and body–brain health may therefore provide an effective framework for understanding and ultimately mitigating age-related cognitive decline.

## 4. LIMITATIONS

A primary limitation of this study is its cross-sectional design, which constrains inference about the temporal dynamics among the examined variables. In addition, participants were not screened for dementia or cognitive impairment, and cognitive status was not used as an exclusion criterion. This raises the possibility that observed age-related effects may partly reflect other age-associated neurological processes or conditions, rather than age alone. To translate these findings into clinical risk assessment, future work will need to validate the proposed models in clinical populations, including individuals with cognitive impairment and dementia. Furthermore, our definitions of physical and lifestyle variables are not exhaustive. To ensure consistency and comparability with research using the UK Biobank, we relied on the variable definitions provided by the study. As a result, some relevant constructs may not be fully captured within our classifications. For example, the UK Biobank categorises intellectual engagement and social interaction as mental health variables rather than lifestyle factors, and such classification decisions may have influenced the observed associations.

Another limitation concerns selection bias within the UK Biobank cohort. The sample is predominantly composed of individuals of White ethnicity, and participants who consented to take part may, on average, be healthier and have greater access to health-related information than the general population, potentially inflating associations with lifestyle and environmental factors ^[172,176,194,195]^. Together, these factors limit the generalisability of the findings. Future studies should include more diverse and underrepresented populations and adopt longitudinal designs to better characterise trajectories of cognitive aging.

Finally, the neuroimaging and cognitive performance measures available in the UK Biobank differ from those commonly used in other large-scale studies. In particular, the neuroimaging data lack cognitively demanding task-based functional MRI (tfMRI) paradigms, which may capture substantial condition-specific variance in certain cognitive domains ^[196]^. Instead, the available tfMRI data are limited in sample size and feature richness and are derived from the Hariri emotional faces task ^[197]^, which primarily probes emotional reactivity and is less cognitively demanding than paradigms such as the *n*-back working memory task or the Face-name paired-associates memory task ^[196,198–200]^. In addition, the UK Biobank cognitive test battery differs from widely used instruments such as the NIH Toolbox for Assessment of Neurological and Behavioral Function ^[201]^ and the Wechsler Adult Intelligence Scale ^[202]^. These differences may further limit the generalisability of cognitive findings across cohorts.

Furthermore, the present analyses focused on a global cognitive composite (*g*-factor), rather than modeling domain-specific cognitive components. While the *g*-factor provides a theoretically motivated summary of shared variance across cognitive measures and is well suited to quantifying individual differences in general cognitive functioning, using it as the primary cognitive outcome may obscure associations that are specific to particular cognitive domains. Alternative factor structures or domain-specific cognitive measures may therefore yield different patterns of associations. Future work should extend the present framework by examining individual cognitive tasks as separate prediction targets to better characterize domain-specific relationships and identify which aspects of cognition are most strongly associated with lifestyle, environmental, body, and brain measures.

Finally, the commonality analysis was conducted using composite lifestyle–environment, body, and brain markers derived from first-level prediction models rather than the original observed variables. This approach was adopted to reduce the high dimensionality and heterogeneity of the three modalities to cognition-relevant predictive summaries that could be compared at the modality level. However, because these composite markers are estimated predictions, they carry sampling variability and model uncertainty, which may attenuate the associations underlying the subsequent commonality analysis and influence the estimated unique and shared components. Accordingly, the reported proportions should be interpreted as overlap among modality-specific cognition-predictive signals, conditional on the performance of the first-level models, rather than as a direct decomposition of variance among the underlying lifestyle, body, and brain variables. Future work using alternative representations of the underlying variables may provide complementary estimates of their relative and shared associations with cognition. Furthermore, because many body and brain measures are strongly associated with chronological age, future age-residualized analyses could help clarify the extent to which their associations with cognition reflect age-independent variation beyond chronological age.

## 5. CONCLUSIONS

In conclusion, our study provides a comprehensive view of how lifestyle and environmental exposures, together with body and brain health, are associated with cognitive functioning in later life. Using machine learning, we extend prior work that has largely focused on isolated systems or within-sample associations ^[9,11,13,20]^ by integrating multiple domains within a single analytical framework. We identify lifestyle and environment domains that may represent potential targets for supporting better cognitive outcomes, providing a foundation for future strategies aimed at improving physical and brain health through behavioral interventions. Importantly, our findings highlight the central role of body and brain health in the relationships between lifestyle, environment, and cognition, underscoring the interconnected influence of behavior, bodily systems, and brain integrity on cognitive aging. Finally, we outline an analytic framework for integrating diverse lifestyle, environmental, body, and brain indicators into multimodal markers of cognition.

While our approach may inform future risk assessment, translational research, and the development of targeted interventions and decision-support tools aimed at supporting cognitive health in later life, the current findings primarily provide a population-level characterization of individual differences in cognitive functioning and generate hypotheses for future research. Further longitudinal research is needed to establish the clinical utility of these markers for individual-level risk assessment, while randomized controlled trials could evaluate whether the lifestyle and environmental factors identified as most informative can be modified to improve cognitive outcomes. The results of this study should also be interpreted in light of the known selection biases in the UK Biobank cohort, including healthy volunteer bias, which may limit the generalizability of the findings to the broader population.

## 6. MATERIALS AND METHODS

We used data from the UK Biobank (Resource Application Number 70132) collected at the baseline assessment and the first imaging visit. UK Biobank has ethical approval from the North West Multi-centre Research Ethics Committee (MREC) as a Research Tissue Bank (RTB; reference 16/NW/0274), and all participants provided informed consent at recruitment.

Cognitive performance and neuroimaging data were obtained during the first imaging visit. For lifestyle/environmental and body physiology measures, we selected the assessment wave that best aligned temporally with the cognitive assessment. When a variable was available at both baseline and the first imaging visit, we used the imaging-visit measure to ensure temporal correspondence with cognitive outcomes. Variables collected only during online or in-person follow-up assessments were taken from the corresponding follow-up wave, and where multiple follow-up waves were available, the wave maximising sample size and feature availability was selected.

Sample sizes varied across analyses as a result of the multimodal, multi-stage modeling framework. Specifically, separate machine learning models were first trained for individual lifestyle and environment domains and for individual body and brain phenotypes, each using the maximum number of participants with complete data for that specific domain or phenotype. Missing values arising from behavioral data processing were retained at this stage (see Data Analysis). Predicted cognitive scores from these domain- and phenotype-specific models were then combined using stacking and entered as explanatory variables in subsequent commonality analyses. Missing values arising from combining predicted values across domains and phenotypes prior to stacking were retained. For the commonality analyses, predicted scores and the observed *g*-factor were matched using participant identifiers, and only individuals with complete data across all explanatory variables included in a given model were used.

Consequently, the final sample size for the commonality analyses was constrained by the smallest domain-specific feature set included at this stage (Category 100108, “Physical activity yesterday”), resulting in an analytical sample of 10,133 participants aged 48–82 years (mean = 64.35, standard deviation, SD = 7.54; 53.97% female; 97.16% White). The initial sample used to derive the general factor of cognition (g-factor) comprised 31,897 participants aged 46–83 years (mean = 64.55, SD = 7.66; 51.32% female; 97.1% White).

### 6.1. Data

#### 6.1.1. General Cognition Factor

Cognitive functioning was operationalized as a latent general cognition factor (*g*-factor) derived from twelve cognitive performance measures spanning eleven tasks: pairs matching, reaction time, numeric memory, prospective memory, tower rearranging, matrix pattern completion, trail making, fluid intelligence, paired associate learning, symbol digit substitution, and picture vocabulary. We used an exploratory structural equation modeling within confirmatory factor analysis framework (ESEM-within-CFA; https://mateuspsi.github.io/esemComp/articles/esem-within-cfa.html) to estimate a hierarchical *g*-factor model (Figure 1), in which the observed cognitive scores load onto intermediate latent factors, which in turn load onto a higher-order *g*-factor.

To evaluate the factor structure, we conducted parallel factor analysis and assessed sampling adequacy using the Kaiser–Meyer–Olkin statistic ^[35]^ and Bartlett’s test of sphericity ^[36]^, confirming that the correlation structure was suitable for factor analysis. Construct validity was then assessed using CFA, with model fit evaluated via the Comparative Fit Index, Tucker– Lewis Index, Root Mean Squared Error of Approximation, Bayesian Information Criterion, and Standardised Root Mean Square Residual ^[37,203,204]^. The cognitive tests included in the model and their corresponding UK Biobank field identifiers are reported in Supplementary Table S1. Factor loadings and goodness-of-fit statistics are provided in Supplementary Tables S2 and S3, respectively.

To reduce skewness in score distributions, time-based measures, including mean reaction time and completion time in the trail making test, were *log*-transformed, and a *log(x+1)* transformation was applied to the number of incorrect matches in the six-pair version of the pairs matching task ^[205,206]^. For the symbol digit substitution test, accuracy was calculated as the proportion of correctly matched symbol–digit pairs relative to the total number attempted, thereby accounting for spurious correct responses arising from rapid guessing and reducing inflation due to chance-level performance. In the prospective memory task, participants were scored as 1 if they correctly selected the target shape (orange circle) on the first attempt and as 0 otherwise (i.e., if a non-target shape was selected). Negative values reflecting invalid or abandoned trials were coded as missing. Zero values were also treated as missing in tasks where a score of zero indicated an invalid observation (e.g., fluid intelligence, picture vocabulary, and trail making duration).

#### 6.1.2. Lifestyle and Environment Domains

Lifestyle and environment variables were represented by twelve domains, including physical activity measured via accelerometry and self-report (daily, yesterday’s, and overall physical activity assessed using metabolic equivalent of task), diet and alcohol consumption, environmental exposures (local environment and sun exposure), sleep quality, smoking, electronic device use, and sexual factors (Figure 1A). Responses coded as “*Do not know*” (−1) or “*Prefer not to answer*” (−3) were retained (73). For categorical variables, these codes were represented as separate categories during one-hot encoding. For numeric or ordinal variables, where one-hot encoding was not applicable, these entries were recoded as missing (NA) but retained in the dataset to allow the modeling algorithm to handle them using its native missing value procedures without imposing arbitrary imputations.

For questionnaire items administered conditionally on a preceding “header” question – an initial item asked of all participants that determined whether follow-up questions were presented – participants were excluded only if they did not respond to the header item itself. Missing values in follow-up questions that were skipped because the header condition was not met were recoded as 0 to indicate that these items were not applicable.

All lifestyle and environment variables and their corresponding UK Biobank field identifiers are listed in Supplementary Table S1.

#### Physical activity: daily activity, yesterday’s activity, metabolic equivalent of task, and accelerometry

Physical activity was assessed using self-report measures and accelerometry.

#### Daily activity

Self-reported physical activity was collected via touchscreen questionnaires and included information on the type and duration of activities undertaken in a typical week, such as walking, do-it-yourself (DIY) activities, moderate and vigorous physical activity, strenuous sports, and time spent in sedentary behaviors, including computer use, television (TV) viewing, and driving. Responses indicating “*Unable to walk*” (−2) for the item “*Number of days/week walked 10+ minutes*” were recoded to 0, as this response is equivalent to reporting zero days of walking. For sedentary behavior items (time spent driving, using a computer, or watching TV), the response code −10 (“*Less than an hour a day*”) was recoded to 0. Categorical items describing types of physical activity, types of transport used, and usual walking pace were one-hot encoded to generate binary indicators for each category.

#### Yesterday’s physical activity

In addition to the weekly measures, time spent in vigorous, moderate, and light physical activity on the previous day was assessed during an online follow-up. Responses were recoded to a 1–7 scale, with higher values indicating greater time spent in each activity type.

#### Metabolic equivalent of task

To quantify activity intensity, metabolic equivalent of task (MET) indices were derived from self-reported physical activity following International Physical Activity Questionnaire (IPAQ) scoring guidelines ^[207]^. A MET represents the ratio of energy expended per kilogram of body weight relative to resting metabolic rate, with 1 MET corresponding to the energy cost of sitting quietly ^[208]^. In the IPAQ framework, MET values assigned to each activity type (walking, moderate, vigorous) are multiplied by activity duration to yield MET-minutes, which are summarised in the UK Biobank as MET-minutes per week. Standard IPAQ MET weights were applied using the following thresholds: walking (3.3 METs), moderate activity (4 METs), and vigorous activity (8 METs) ^[208]^. MET-minutes per week were available separately for walking, moderate, and vigorous activity, as well as summed across all activity types ^[207]^. Participants were additionally classified into IPAQ activity groups (low, moderate, high) and assigned binary indicators reflecting whether they met IPAQ recommendations for moderate/vigorous or moderate/vigorous/walking activity. Summary measures of total activity days and total activity minutes were also included.

#### Accelerometry

Instrumental measures of physical activity were obtained from accelerometry data collected from 103,567 participants over seven days using an Axivity AX3 wrist-worn triaxial accelerometer. Acceleration was recorded in milli-gravities in 5-second epochs, yielding 120,960 data points per participant ^[60]^. Analyses included mean daily, hourly, and overall acceleration; acceleration intensity distributions reflecting the proportion of time spent at or below specified acceleration thresholds; accelerometer wear time; and derived measures indexing the average proportion of time spent in light and moderate-to-vigorous physical activity, as well as sedentary behavior and sleep ^[209]^.

#### Environmental exposures: local environment and sun exposure

##### Local environment

Local environmental characteristics included residential air pollution estimates for 2005– 2007 and 2010 (e.g., distance to major roads and traffic load on the nearest road) ^[210]^, residential noise pollution (16-hour, 24-hour, daytime, evening, and night-time noise levels), and indicators of greenspace and coastal proximity. Greenspace metrics included the percentage of greenspace, domestic garden coverage, water coverage, and the proportion of natural versus built environment within 300 m and 1000 m buffers. Water quality indicators included mineral content and water hardness, defined according to United States Geological Survey and World Health Organization standards ^[211]^.

##### Sun exposure

Sun exposure was assessed via touchscreen questionnaire and included time spent outdoors in summer and winter, skin color, ease of tanning, childhood sunburn frequency, natural hair color, facial aging, use of sun/ultraviolet (UV) protection, and frequency of solarium or sunlamp use. For time spent outdoors in summer and winter and solarium/sunlamp use, responses coded as −10 (“*Less than 1”*) were recoded to 0.5. Use of sun/UV protection was set to 0 for participants who reported never being outdoors in sunshine (code 5). Skin color, hair color, and facial aging were treated as categorical variables and one-hot encoded.

##### Electronic device use

Electronic device use included mobile phone use (duration and frequency), hands-free or speakerphone use, changes in mobile phone use over the previous two years, preferred side of the head during calls, and engagement in computer gaming. This variable reflects overall electronic device use, without distinguishing between leisure and work-related use. Variables reflecting change in phone use over time and preferred head side were treated as categorical and one-hot encoded.

##### Sexual factors

Sexual factors were derived from touchscreen questionnaire items capturing age at first sexual intercourse, lifetime number of sexual partners (including same-sex partners), and related behavioral indicators. For participants who reported never having had sex (code −2), missing values in follow-up items (e.g., lifetime number of sexual partners, ever having had same-sex intercourse) were recoded to 0 to indicate non-applicability. Similarly, for participants who reported never having had same-sex intercourse, missing values for the lifetime number of same-sex partners were set to 0. Age at first sexual intercourse was grouped into three categories: “*Never had sex*” (−2), “*Do not know/Prefer not to answer*” (−1/−3), and valid ages. Non-response categories were one-hot encoded to retain information without imposing numeric meaning. The original continuous age variable was retained, with non-response codes recoded as missing to ensure the variable reflected only valid age values.

##### Sleep

Sleep characteristics were assessed using touchscreen questionnaire items on sleep duration, daytime napping, unintentional dozing, snoring, insomnia/sleeplessness, and chronotype (morning/evening preference). Chronotype was treated as categorical and one-hot encoded. Snoring was recoded as a binary variable (Yes = 1, No = 0).

##### Diet

Diet was assessed using touchscreen questionnaire items capturing the frequency of consumption of common food and drink categories, including fruits and vegetables, meat and fish, eggs, dairy products, bread and cereals, tea and coffee, water intake, salt and sugar use, and overall dietary variation. Several follow-up items were conditional: cereal type was recorded only for participants who reported eating at least one bowl of cereal per week; coffee type only for those who drank coffee at least occasionally; bread type for participants who consumed at least one slice of bread per week; cheese intake for those consuming dairy products; and non-butter spread type for participants who reported using spreads other than butter or spreadable butter.

For frequency variables, responses coded as −10 (“*Less than one*”) were recoded to 0.5. Categorical indicators for non-consumption or product type (e.g., never eat eggs, dairy, wheat, or sugar; milk type; spread type; bread type; cereal type; coffee type; major dietary changes) were one-hot encoded. Hot drink temperature was recoded to an ordered scale (Very hot = 3; Hot = 2; Warm = 1; Do not drink hot drinks = 0). Responses to the “Age when last ate meat” item were excluded due to limited coverage.

##### Alcohol consumption

Alcohol consumption was assessed via touchscreen questionnaire items covering drinking status, frequency, beverage type, whether alcohol was usually consumed with meals, and, for former drinkers, reasons for cessation and time since stopping. Participants who reported never drinking alcohol had all alcohol-related variables set to 0. Weekly intake measures were collected from participants drinking more than once or twice per week; monthly intake was recorded for those drinking one to three times per month or only on special occasions. Missing values for follow-up items outside a participant’s relevant intake category were set to 0. Similarly, variables capturing usual alcohol consumption with meals, reasons for reducing intake, or reasons for stopping alcohol were set to 0 when not applicable.

Alcohol intake frequency (Daily/almost daily = 5; Three or four times a week = 4; Once or twice a week = 3; One to three times a month = 2; Special occasions only = 1; Never = 0) and hot drink temperature (Very hot = 3; Hot = 2; Warm = 1; Do not drink hot drinks = 0) were recoded to ordered scales. For the item “Alcohol usually taken with meals,” the –6 (“It varies”) response was recoded to 0.5 (Yes = 1, No = 0, It varies = 0.5). All categorical alcohol variables, including drinker status, change over the past ten years, and reasons for reducing or stopping drinking, were one-hot encoded.

##### Smoking

Smoking behavior was assessed via touchscreen questionnaire items covering smoking status, duration, number of cigarettes per day, age at initiation, and, for former smokers, amount previously smoked, time since cessation, ease of quitting, and reasons for stopping. Exposure to environmental tobacco smoke inside and outside the home was also recorded. Participants who did not respond to the “*Ever smoked*” item were excluded. Responses coded as −10 (“*Less than one*”) for the number of cigarettes currently or previously smoked per day were recoded to 0.5.

Follow-up items were handled according to questionnaire routing. Variables asked only of current smokers (e.g., age started smoking, light smoking, type of tobacco currently smoked, current cigarette count, nicotine-dependence indicators, and smoking-reduction questions) were set to 0 for participants who did not report current smoking. Variables asked only of former smokers (e.g., age stopped smoking, age started smoking, previous cigarette count, type of tobacco previously smoked, cessation-history items) were set to 0 for participants who did not report past smoking. Items for specific subgroups (e.g., cigar or pipe smokers with previous cigarette use) were set to 0 for ineligible participants. All categorical smoking variables were mapped to descriptive categories and one-hot encoded. Multi-select questions were collapsed so that each category was represented by a single binary indicator.

#### 6.1.3. Body phenotypes

We built bodily markers of cognition from 19 body phenotype groups, capturing the function and structure of nine systems: cardiovascular, pulmonary, renal, hepatic, immune, metabolic, musculoskeletal, sensory, and body/abdominal organ composition, derived from Categories 100006 (“Physical measures”) and 100003 (Imaging). To maintain a harmonised and comparable feature set across domains, we restricted analyses to variables available within these two core UK Biobank categories. Phenotype-level markers were then combined into a composite body marker using a multimodal stacking approach (see Data Analysis section) ^[21,132]^.

##### Cardiovascular and pulmonary systems

Cardiovascular and pulmonary systems were assessed using six phenotypes: carotid ultrasound, arterial stiffness indices, pulse wave analysis, heart MRI, 12-lead resting electrocardiogram (ECG), and cardiopulmonary function measures, including forced vital capacity (FVC), forced expiratory volume in 1 second (FEV_1_), peak expiratory flow, pulse rate, and blood pressure.

##### Renal and hepatic systems

Renal and hepatic systems were evaluated using MRI-derived structural measures and blood/urine assays, including glomerular filtration, urine electrolytes, creatinine, and blood levels of albumin, urea, urate, cystatin C, phosphate, total protein, alanine and aspartate aminotransferases, γ-glutamyl transferase, bilirubin, and alkaline phosphatase. These measures were summarised into three phenotypes: liver MRI, kidney MRI, and renal/hepatic blood markers.

##### Immune system

Immune function was captured through blood analysis, including C-reactive protein, haemoglobin, and leukocyte, erythrocyte, and thrombocyte counts.

##### Metabolism

Metabolism was assessed via blood markers of glucose and lipid metabolism (apolipoproteins A and B, cholesterol, glucose, glycated haemoglobin, high-density lipoprotein cholesterol (HDL-C), low-density lipoprotein cholesterol (LDL-C), lipoprotein A, triglycerides) and by circulating levels of calcium, vitamin D, insulin-like growth factor-1 (IGF-1), and testosterone, which could not be grouped elsewhere.

##### Musculoskeletal system

Musculoskeletal health was evaluated using three phenotypes: bone size, mineral content, and density measured with dual-energy X-ray absorptiometry (DXA), heel bone density measured via quantitative ultrasound, and anthropometric measures including handgrip strength, height, weight, body mass index, waist and hip circumference, and ankle spacing width.

##### Body composition

Body composition was assessed using MRI, bioelectrical impedance, and DXA, while abdominal organ composition was measured via MRI, yielding four phenotypes: body composition by impedance, body composition by DXA, abdominal composition by MRI, and abdominal organ composition by MRI.

##### Sensory system

The sensory system was evaluated using the speech-reception threshold, defined as the signal-to-noise ratio at which participants correctly understood 50% of the presented speech.

For variables measured on both sides of the body, specifically handgrip strength, heel bone mineral density, and ankle spacing width, or repeated within a visit (pulse rate, diastolic and systolic blood pressure), values were averaged. For the augmentation index from pulse wave analysis, the first measurement was used. Spirometry measures, including FVC, FEV_1_, and peak expiratory flow, were derived from the best performance trial, and additional ratios, such as FVC/FEV_1_ and waist-to-hip circumference, were computed manually. Measures with >60% missing data (rheumatoid factor, blood oestradiol, urine microalbumin, cardiorespiratory fitness, exercise ECG, kidney distance, total adipose tissue volume, and total lean tissue volume) were excluded ^[23,212]^. Measures included in each body physiology phenotype, together with the corresponding UK Biobank field numbers for the original variables and the derived measures used in the analyses, are listed in Supplementary Table S1.

#### 6.1.4. Brain Phenotypes

We derived brain markers of cognition from 81 neuroimaging phenotypes spanning three modalities: diffusion-weighted MRI (dwMRI), resting-state functional MRI (rsMRI), and structural MRI (sMRI). MRI acquisition protocols are available at http://biobank.ctsu.ox.ac.uk/crystal/refer.cgi?id=2367, and processing pipelines are described in the UK Biobank brain imaging documentation (https://biobank.ctsu.ox.ac.uk/crystal/crystal/docs/brain_mri.pdf) ^[213,214]^. Phenotype-specific markers were then combined within and across modalities to obtain unimodal and composite brain markers.

Prior to predictive modeling, neuroimaging phenotypes were adjusted for common and modality-specific confounds ^[215]^. Confounds with pairwise correlations exceeding |*r*| = 0.7 were removed to reduce multicollinearity ^[132]^. A full list of neuroimaging confounds is provided in Supplementary Table S1.

##### Diffusion-weighted MRI (dwMRI)

dwMRI included 42 phenotypes, comprising UK Biobank imaging-derived phenotypes ^[214]^ and structural connectomes reconstructed by Mansour and colleagues ^[216]^. Voxelwise microstructural measures were obtained using diffusion tensor imaging (DTI: fractional anisotropy (FA), diffusion tensor mode, mean diffusivity, and tensor eigenvalues) and neurite orientation dispersion and density imaging (NODDI: intracellular volume fraction, isotropic/free water fraction, orientation dispersion index). These were summarised for 48 white matter tracts using tract-based spatial statistics and 27 tracts via probabilistic tractography. Structural connectomes were reconstructed using multi-shell, multi-tissue constrained spherical deconvolution, followed by whole-brain probabilistic tractography ^[216]^. Tractography streamlines were combined with six cortical–subcortical atlas combinations to generate region-to-region connectivity matrices, capturing streamline count, fibre bundle capacity, mean streamline length, and mean FA. The six atlas combinations included: (1) the Desikan–Killiany cortical atlas ^[217]^ with Melbourne Subcortical Atlas scale I (MSA-I) ^[218]^, (2) the Destrieux cortical atlas ^[219]^ with MSA-I ^[218]^, (3) the Glasser cortical atlas ^[220]^ with MSA-I ^[218]^, (4) the Glasser cortical atlas ^[220]^ with MSA-IV ^[218]^, (5) the Schaefer atlas for 200 cortical regions and 7 networks ^[221,222]^ with MSA-I ^[218]^, and (6) the Schaefer atlas for 500 cortical regions and 7 networks ^[221,222]^ with MSA-IV ^[218]^.

##### Resting-state functional MRI (rsMRI)

rsMRI included 18 phenotypes capturing functional connectivity between large-scale networks ^[214]^ and parcellated nodes ^[216]^. Network-level phenotypes were derived via independent component analysis (ICA) and included full and partial correlation matrices among 21 and 55 networks, as well as network amplitude measures. Parcellation-based functional connectomes used the same six atlas combinations as dwMRI, with full and partial correlation matrices computed from blood-oxygen-level-dependent (BOLD) time series using Nilearn’s *ConnectivityMeasure* function ^[132]^.

##### Structural MRI (sMRI)

sMRI included 21 phenotypes derived from T1- and T2-weighted images, encompassing subcortical volumes and intensities, cortical and white matter morphometrics (surface area, thickness, volume), grey–white matter contrast, pial surface area, and total white matter hyperintensity volume. These measures were extracted using FSL FAST, FSL FIRST, FreeSurfer ASEG and subcortical volumetric subsegmentation pipelines, ex vivo Brodmann area maps, and the Destrieux, Desikan–Killiany–Tourville, and Desikan–Killiany parcellations^[132]^.

#### 6.1.5. Data Analysis Machine learning

To build predictive markers of cognition, we employed a two-stage multimodal stacking framework. In the first stage, a series of XGBoost models ^[223]^ were trained to predict the *g*-factor from each lifestyle and environment domain, and from each body or brain phenotype. These first-level models captured domain- and phenotype-specific associations with cognition. In the second stage, outputs from the first-level models served as input features for a second-level integrative model, implemented using a Random Forest algorithm ^[224–226]^. Missing values arising from combining phenotype- and domain-specific predictions were retained in the input features. Seven stacked models were constructed: (1) a composite lifestyle–environment marker integrating all twelve domains, (2) a composite body marker integrating all 19 body phenotypes, (3) a composite brain marker integrating all 81 neuroimaging phenotypes, (4–6) modality-specific brain models for dwMRI, rsMRI, and sMRI, and (7) a body–brain model integrating both body and brain phenotypes.

For model training and evaluation, we employed nested cross-validation to minimise bias and prevent data leakage. Five outer folds were used for performance estimation, with each iteration holding out 20% of the data as an independent test set and using the remaining 80% for training. Within each outer-fold training set, hyperparameter optimization was performed using ten inner cross-validation folds, with models trained on nine partitions and validated on the remaining partition, cycling through all fold combinations. For each outer fold, we selected the hyperparameter configuration yielding the lowest mean squared error (*MSE*) across inner folds and then retrained the model on the full outer-fold training set before evaluation on the held-out test set. This process was repeated across all five outer folds to ensure each participant contributed to both training and testing phases, but never within the same fold. Hyperparameter tuning included up to twelve parameters for XGBoost and two parameters for Random Forest (see parameter grid for each algorithm in Supplementary Table S12). For analytic samples exceeding 30,000 features, hyperparameter optimization was performed using randomised search (*RandomizedSearchCV*), and for smaller feature sets, a full grid search (*GridSearchCV*) was applied.

Throughout the analysis, models were trained exclusively on outer-fold training data and evaluated only on the associated test data. Predictive performance was quantified using Pearson correlation (*r*), coefficient of determination (*R*²), mean absolute error (*MAE*), and *MSE* between predicted and observed *g*-factors.

##### Data standardization

All features were standardized within each outer-fold training set, and the resulting mean and standard deviation were applied to the corresponding test set. Standardization was applied at multiple stages: cognitive scores were standardized prior to ESEM; neuroimaging measures were standardized before deconfounding; lifestyle, environment, and body physiology features were standardized before first-level model training; and both first-level predictions and observed *g*-factors were standardized before training stacked models. This procedure ensured consistent feature scaling while preventing data leakage.

##### Statistical significance

Model significance was evaluated using a bootstrap procedure applied to predicted and observed *g*-factors pooled across all five outer-fold test sets. An empirical distribution of Pearson correlations was generated via 5,000 resampling iterations, with 95% confidence intervals (CIs) derived from these distributions. Performance was considered statistically significant if the CI did not include zero.

We also benchmarked predictive of the composite lifestyle–environment marker against individual lifestyle and environment domains, individual body and brain phenotypes, unimodal brain markers, the composite brain marker, the composite body marker, and the body–brain marker. Differences in performance were quantified using *r*, *R*², *MSE*, and *MAE*.

##### Feature importance

Feature importance was assessed using the Haufe transformation ^[39,227–229]^, which maps model outputs back to the original feature space while accounting for the data covariance structure, facilitating interpretation of associations between input features and predictions. Domain-level importance was quantified by correlating predictions from each individual-domain model with the composite lifestyle–environment marker. Feature-level importance was calculated by correlating each feature with the composite marker. To confirm the directionality of feature weights derived from the Haufe transformation, we additionally computed correlations between each feature and the observed *g*-factor. To account for multiple comparisons, we applied Bonferroni correction.

#### 6.1.6. Commonality Analysis

We employed commonality analysis ^[230,231]^ to quantify (a) the proportion of the association between lifestyle and environment and cognition explained by body and brain markers, and (b) the extent to which composite lifestyle–environment and body–brain markers jointly account for age-related variation in cognition.

##### Two-predictor framework

We first implemented a two-predictor framework in which the observed *g*-factor served as the response variable, and the composite lifestyle–environment and body and/or brain markers, pooled across the five outer-fold test sets, served as explanatory variables in linear regression models:

1. **Model 1:** *g*_observed_ ∼ *ĝ*_lifestyle–environment_
2. **Model 2:** *g*_observed_ ∼ *ĝ*_brain and/or body_
3. **Model 3:** *g*_observed_ ∼ *ĝ*_lifestyle–environment_ + *ĝ*_brain and/or body_

From these models, we estimated the total variance explained (*R*^2^) and decomposed it into unique (attributable to each explanatory variable) and common (jointly explained by both explanatory variables) variance components ^[230,231]^:

##### Unique variance components

- **Unique_lifestyle–environment_** = *R*^2^_lifestyle–environment,body and/or brain_ – *R*^2^_body and/or brain_
- **Unique_body and/or brain_** = *R*^2^_lifestyle–environment,body and/or brain_ – *R*^2^_lifestyle–environment_

### Common variance component

- **Common_lifestyle–environment, body and/or brain_** = *R*^2^_lifestyle–environment,body and/or brain_ – Unique_lifestyle–environment_ – Unique_body and/or brain_ = *R*^2^_lifestyle–environment_ + *R*^2^_body and/or brain_ – *R*^2^_lifestyle–environment,body and/or brain_

The proportion of the lifestyle–environment–cognition association explained by body and/or brain markers was expressed as a percentage, defined as the ratio of the jointly explained variance to the total variance explained by the lifestyle–environment marker:

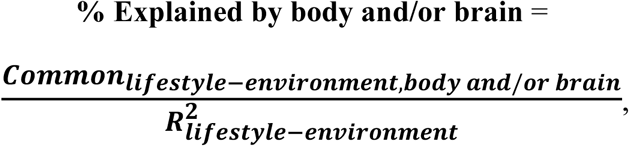

where *R*^2^_lifestyle–environment_ = Common_lifestyle–environment,body and/or brain_ + Unique_lifestyle–environment_.

#### Three-predictor framework

To evaluate the proportion of age-related cognitive variance explained uniquely and jointly by lifestyle–environment and body–brain markers, we extended the analysis to include age:

1. **Model 1:** *g*_observed_ ∼ *ĝ*_lifestyle–environment_
2. **Model 2**: *g*_observed_ ∼ *ĝ*_body–brain_
3. **Model 3:** *g*_observed_ ∼ age
4. **Model 4:** *g*_observed_ ∼ *ĝ*_lifestyle–environment_ + *ĝ*_body–brain_ + age

Similarly, we decomposed the total variance (*R*^2^) from Model 4 into components uniquely explained by age, lifestyle–environment, and body–brain markers, as well as variance jointly explained by each pair of predictors and by all three predictors together.

#### Unique variance components

- **Unique_age_** = *R*^2^_age,lifestyle–environment,body–brain_ – *R*^2^_lifestyle–environment,body–brain_
- **Unique_lifestyle–environment_** = *R*^2^_age,lifestyle–environment, body–brain_ – *R*^2^_age,body–brain_
- **Unique_body–brain_** = *R*^2^_age,lifestyle–environment,body–brain_ – *R*^2^_age,lifestyle–environment_

#### Common variance components

- **Common_age,lifestyle–environment_** = *R*^2^_age,body–brain_ + *R*^2^_lifestyle–environment,body–brain_ – *R*^2^_body– brain_ – *R*^2^_age,lifestyle–environment,body–brain_
- **Common_age,body–brain_** = *R*^2^_age,lifestyle–environment_ + *R*^2^_lifestyle–environment,body–brain_ – *R*^2^_lifestyle– environment_ – *R*^2^_age,lifestyle–environment,body–brain_
- **Common_lifestyle–environment,body–brain_** = *R*^2^_age,lifestyle–environment_ + *R*^2^_age,body–brain_ – *R*^2^_age_ – *R*^2^_age,lifestyle–environment,body–brain_
- **Common_age, lifestyle–environment, body–brain_** = *R*^2^_age_ + *R*^2^_lifestyle–environment_ + *R*^2^_body–brain_ – *R*^2^_age,lifestyle–environment_ – *R*^2^_age,body–brain_ – *R*^2^_lifestyle–environment,body–brain_ + *R*^2^_age,lifestyle–environment,body–brain_

To quantify the proportion of age-related cognitive variance explained by each marker alone and in combination, we calculated percentage ratios. The numerator represented the variance in the observed *g*-factor jointly explained by age and the marker(s), and the denominator represented the total variance explained by age.

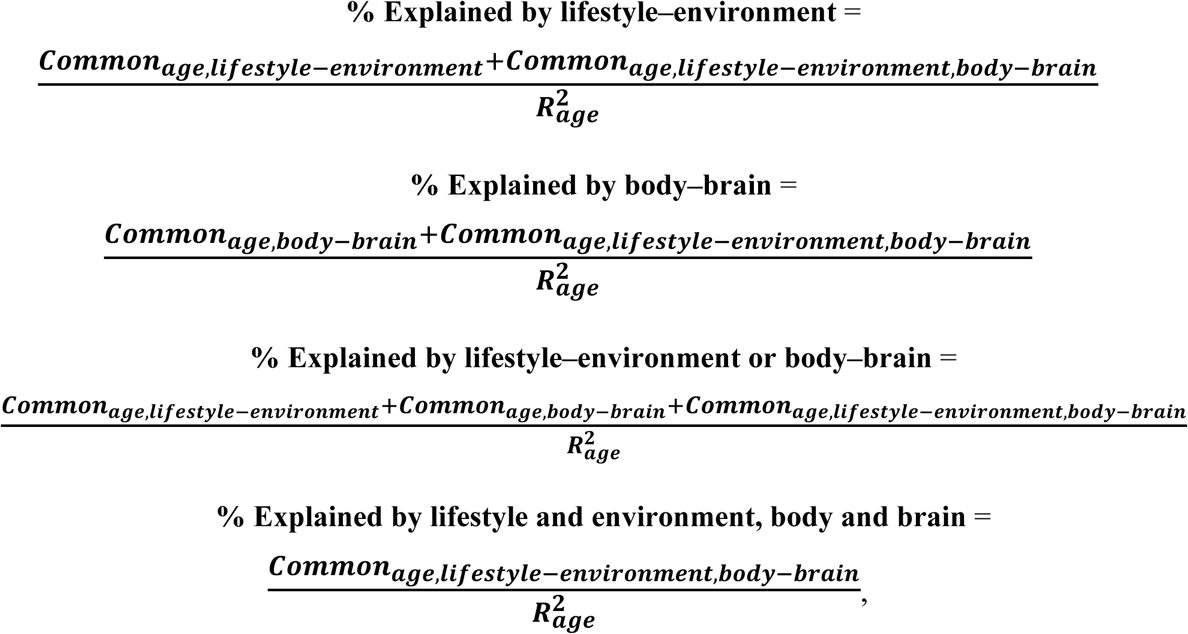

where *R*^2^_age_ = Unique_age_ + Common_age,lifestyle–environment_ + Common_age,body–brain_ + Common_age,lifestyle–environment,body–brain_.

## Data Availability

The data used in this study are subject to restrictions. Access requests should be directed to the UK Biobank (https://www.ukbiobank.ac.uk/). This manuscript is a computational study and did not generate any new data. Analyses were conducted using the UK Biobank Resource under Application Number 70132. The modelling code is available on GitHub.

https://github.com/HAM-lab-Otago-University/UKBiobank-Lifestyle-Environment

## 7. Author Contributions

IB: Conceptualization, Methodology, Software, Formal analysis, Investigation, Data Curation, Writing – Original Draft, Writing – Review & Editing, Visualization; NP: Conceptualization, Methodology, Writing – Review & Editing, Resources, Supervision, Project administration, Funding acquisition.

## 8. Acknowledgements

This research has been conducted using the UK Biobank Resource under Application Number 70132. N.P. was supported by the Health Research Council of New Zealand (grant numbers 21/618 and 24/838), by the Neurological Foundation of New Zealand (grant number 2350 PRG), and by the Ministry of Business, Innovation and Employment (grant numbers UOA2421 and RTVU2403). I.B. was supported by the University of Otago.

AI tools, including ChatGPT and Copilot, were used solely for language editing and did not contribute to the study design, data analysis, interpretation of results, or generation of scientific content. All analyses, results, interpretations, and scientific conclusions were produced by the authors based on the original study data and analyses.

## 10. Data Availability

The data used in this study are subject to access restrictions and are available through the UK Biobank (https://www.ukbiobank.ac.uk/) upon approved application. This study is computational in nature and did not generate new data. The research was conducted under UK Biobank Resource Application Number 70132.

## 11. Code Availability

All modeling code is available on GitHub at https://github.com/irinabuianova/UKBiobank-Lifestyle-Environment.

## References

[1] H. Schickedanz, C. Safaeipour, D. Sherzai, A. Sherzai, Am. J. Lifestyle Med. 2026, 15598276261417271.

[2] A. N. Vidyanti, F. Rahmawati, R. H. Rahman, A. Prodjohardjono, A. Gofir, J. Prev. Alzheimers Dis. 2025, 12, 100028.

[3] J. Wang, C. Chen, J. Zhou, Z. Xu, L. Xu, X. Li, Z. Zhong, Y. Lv, X. Shi, Nat. Commun. 2025, 16, 539.

[4] M. N. Key, A. N. Szabo-Reed, Nutrients 2023, 15, 2495.

[5] K. R. Sewell, K. I. Erickson, S. R. Rainey-Smith, J. J. Peiffer, H. R. Sohrabi, B. M. Brown, Neurosci. Biobehav. Rev. 2021, 130, 369.

[6] J. W. Pickersgill, C. V. Turco, K. Ramdeo, R. S. Rehsi, S. D. Foglia, A. J. Nelson, Front. Psychol. 2022, 13, DOI 10.3389/fpsyg.2022.831819.

[7] N. Wang, T. Li, Sage Open 2025, 15, 21582440251386694.

[8] M. Zarringhadam, S. Hasanvand, M. Birjandi, A. Beiranvand, BMC Res. Notes 2024, 17, 101.

[9] W. Y. Tan, C. A. Hargreaves, N. Kandiah, S. Hilal, J. Prev. Alzheimers Dis. 2024, 11, 13.

[10] K. X. Ye, L. Sun, L. Wang, A. L. Y. Khoo, K. X. Lim, G. Lu, L. Yu, C. Li, A. B. Maier, L. Feng, Neurosci. Biobehav. Rev. 2023, 152, 105286.

[11] T. Campbell, B. Cullen, Int. J. Epidemiol. 2023, 52, 1592.

[12] E. S. Nichols, G. Nelson, C. J. Wild, A. M. Owen, PLOS ONE 2024, 19, e0298899.

[13] M. Wen, H. Zheng, J.-H. Chen, T. C. T. Fong, Y. Li, D. Su, Humanit. Soc. Sci. Commun. 2025, 12, 214.

[14] A. T. Aborode, O. A. Emmanuel, I. A. Onifade, E. Olotu, O. J. Otorkpa, Q. Mehmood, S. I. Abdulai, A. Jamiu, A. Osinuga, C. I. Oko, S. Fakorede, M. Mangdow, O. Babatunde, Z. Olapade, A. G. Victoria, A. Salami, I. A. Usman, V. I. Agboli, R. O. Adesola, NeuroMarkers 2025, 2, 100034.

[15] S. Espinosa-Salas, M. Gonzalez-Arias, in StatPearls, StatPearls Publishing, Treasure Island (FL), 2025.

[16] J. M. Rippe, Am. J. Lifestyle Med. 2018, 12, 499.

[17] J. A. Potashkin, D. J. Vidyadhara, H. C. Hunsberger, Am. J. Lifestyle Med. 2025, 15598276251411888.

[18] J. Mintzer, K. A. Donovan, A. Z. Kindy, S. L. Lock, L. R. Chura, N. Barracca, Front. Med. 2019, 6, 204.

[19] E. K. Hui, A. Sommerlad, H. Naismith, Lancet Healthy Longev. 2025, 6, DOI 10.1016/j.lanhl.2025.100735.

[20] M. Bloomberg, G. Muniz-Terrera, L. Brocklebank, A. Steptoe, Nat. Commun. 2024, 15, 5003.

[21] Y. E. Tian, J. H. Cole, E. T. Bullmore, A. Zalesky, Nat. Ment. Health 2024, 2, 1250.

[22] N. R. DeJong, J. F. A. Jansen, M. P. J. van Boxtel, M. T. Schram, C. D. A. Stehouwer, M. M. J. van Greevenbroek, C. J. H. van der Kallen, A. Koster, S. J. P. M. Eussen, B. E. de Galan, W. H. Backes, S. Köhler, Brain Commun. 2024, 6, fcae171.

[23] I. Buianova, N. Pat, Npj Aging 2026, 12, 108.

[24] C. Pierpaoli, P. Jezzard, P. J. Basser, A. Barnett, G. Di Chiro, Radiology 1996, 201, 637.

[25] M. H. Lee, C. D. Smyser, J. S. Shimony, AJNR Am. J. Neuroradiol. 2013, 34, 1866.

[26] M. Symms, H. R. Jäger, K. Schmierer, T. A. Yousry, J. Neurol. Neurosurg. Psychiatry 2004, 75, 1235.

[27] C. Krämer, J. Stumme, L. da Costa Campos, P. Dellani, C. Rubbert, J. Caspers, S. Caspers, C. Jockwitz, GeroScience 2024, 46, 283.

[28] E. Dhamala, K. W. Jamison, A. Jaywant, S. Dennis, A. Kuceyeski, Hum. Brain Mapp. 2021, 42, 3102.

[29] J. Rasero, A. I. Sentis, F.-C. Yeh, T. Verstynen, PLOS Comput. Biol. 2021, 17, e1008347.

[30] C. Sripada, S. Rutherford, M. Angstadt, W. K. Thompson, M. Luciana, A. Weigard, L. H. Hyde, M. Heitzeg, Mol. Psychiatry 2020, 25, 3413.

[31] Z. Jost, S. Kujach, Clin. Interv. Aging 2025, 20, 459.

[32] Y. Y. Hoogendam, A. Hofman, J. N. van der Geest, A. van der Lugt, M. A. Ikram, Eur. J. Epidemiol. 2014, 29, 133.

[33] T. A. Salthouse, Intelligence 2013, 41, 501.

[34] A. R. Jensen, Novartis Found. Symp. 2000, 233, 37.

[35] H. F. Kaiser, Psychometrika 1974, 39, 31.

[36] M. S. Bartlett, J. R. Stat. Soc. Ser. B Methodol. 1954, 16, 296.

[37] Y. Xia, Y. Yang, Behav. Res. Methods 2019, 51, 409.

[38] C. M. Williams, G. Labouret, T. Wolfram, H. Peyre, F. Ramus, Behav. Genet. 2023, 53, 85.

[39] S. Haufe, F. Meinecke, K. Görgen, S. Dähne, J.-D. Haynes, B. Blankertz, F. Bießmann, NeuroImage 2014, 87, 96.

[40] Y. Lee, J. Kim, J. H. Back, Prev. Med. 2009, 48, 86.

[41] H. J. Lee, J. Jang, D.-W. Choi, W. Chae, E.-C. Park, S.-I. Jang, BMC Geriatr. 2020, 20, 317.

[42] L. Clare, Y.-T. Wu, J. C. Teale, C. MacLeod, F. Matthews, C. Brayne, B. Woods, C.-W. study Team, PLOS Med. 2017, 14, e1002259.

[43] R. Eramudugolla, M. H. Huque, J. Wood, K. J. Anstey, J. Am. Med. Dir. Assoc. 2021, 22, 399.

[44] L. Eudave, M. A. Pastor, Aging 2023, 15, 887.

[45] S. Depestele, V. Ross, S. Verstraelen, K. Brijs, T. Brijs, K. van Dun, R. Meesen, Transp. Res. Part F Traffic Psychol. Behav. 2020, 73, 433.

[46] L. Chen, D. B. Carr, R. K. Singh, S. Bekena, Y. Zhu, K. Taylor, J.-F. Trani, G. M. Babulal, Neurology 2025, 105, e214440.

[47] M. A. Hird, P. Egeto, C. E. Fischer, G. Naglie, T. A. Schweizer, J. Alzheimers Dis. JAD 2016, 53, 713.

[48] H. Wang, H. Zhang, Y. Chen, M. Cai, C. Guo, P. Chen, Front. Aging Neurosci. 2022, 14, 935291.

[49] T. Skillbäck, K. Blennow, H. Zetterberg, J. Skoog, L. Rydén, H. Wetterberg, X. Guo, S. Sacuiu, M. M. Mielke, A. Zettergren, I. Skoog, S. Kern, Alzheimers Dement. 2022, 18, 1667.

[50] N. del Campo, P. Payoux, A. Djilali, J. Delrieu, E. O. Hoogendijk, Y. Rolland, M. Cesari, M. W. Weiner, S. Andrieu, B. Vellas, For the MAPT/DSA Study Group, Neurology 2016, 86, 36.

[51] L. L. Y. Chan, M. T. Espinoza Cerda, M. A. Brodie, S. R. Lord, M. E. Taylor, Int. Psychogeriatr. 2025, 37, 100031.

[52] M. M. Gonzales, C.-P. Wang, M. Quiben, D. MacCarthy, S. Seshadri, M. Jacob, H. Hazuda, Int. J. Geriatr. Psychiatry 2020, 35, 897.

[53] Z. Meiner, E. Ayers, J. Verghese, Ann. Geriatr. Med. Res. 2020, 24, 3.

[54] V. N. Poole, S. Oveisgharan, L. Yu, R. J. Dawe, S. E. Leurgans, S. Zhang, K. Arfanakis, A. S. Buchman, D. A. Bennett, Front. Aging Neurosci. 2023, 15, 1194986.

[55] A. Ezzati, M. J. Katz, M. L. Lipton, R. B. Lipton, J. Verghese, Neuroradiology 2015, 57, 851.

[56] A. M. V. Wennberg, R. Savica, M. M. Mielke, Dement. Geriatr. Cogn. Disord. 2017, 43, 128.

[57] O. Hansson, M. Svensson, A.-M. Gustavsson, E. Andersson, Y. Yang, K. Nägga, U. Hållmarker, S. James, T. Deierborg, Alzheimers Res. Ther. 2019, 11, 87.

[58] K. I. Erickson, C. Hillman, C. M. Stillman, R. M. Ballard, B. Bloodgood, D. E. Conroy, R. Macko, D. X. Marquez, S. J. Petruzzello, K. E. Powell, Med. Sci. Sports Exerc. 2019, 51, 1242.

[59] Y. C. Klimentidis, D. A. Raichlen, J. Bea, D. O. Garcia, N. E. Wineinger, L. J. Mandarino, G. E. Alexander, Z. Chen, S. B. Going, Int. J. Obes. 2005 2018, 42, 1161.

[60] A. Doherty, D. Jackson, N. Hammerla, T. Plötz, P. Olivier, M. H. Granat, T. White, V. T. van Hees, M. I. Trenell, C. G. Owen, S. J. Preece, R. Gillions, S. Sheard, T. Peakman, S. Brage, N. J. Wareham, PloS One 2017, 12, e0169649.

[61] K. Yaffe, T. D. Hoang, A. L. Byers, D. E. Barnes, K. E. Friedl, Alzheimers Dement. 2014, 10, S111.

[62] M. Cefis, R. Chaney, J. Wirtz, A. Méloux, A. Quirié, C. Leger, A. Prigent-Tessier, P. Garnier, Front. Mol. Neurosci. 2023, 16, DOI 10.3389/fnmol.2023.1275924.

[63] A. M. Stein, T. M. V. Silva, F. G. de M. Coelho, F. J. Arantes, J. L. R. Costa, E. Teodoro, R. F. Santos-Galduróz, Dement. Neuropsychol. 2018, 12, 114.

[64] C. D. Wrann, J. P. White, J. Salogiannnis, D. Laznik-Bogoslavski, J. Wu, D. Ma, J. D. Lin, M. E. Greenberg, B. M. Spiegelman, Cell Metab. 2013, 18, 649.

[65] M. V. Lourenco, R. L. Frozza, G. B. de Freitas, H. Zhang, G. C. Kincheski, F. C. Ribeiro, R. A. Gonçalves, J. R. Clarke, D. Beckman, A. Staniszewski, H. Berman, L. A. Guerra, L. Forny-Germano, S. Meier, D. M. Wilcock, J. M. de Souza, S. Alves-Leon, V. F. Prado, M. A. M. Prado, J. F. Abisambra, F. Tovar-Moll, P. Mattos, O. Arancio, S. T. Ferreira, F. G. De Felice, Nat. Med. 2019, 25, 165.

[66] L. Mandolesi, A. Polverino, S. Montuori, F. Foti, G. Ferraioli, P. Sorrentino, G. Sorrentino, Front. Psychol. 2018, 9, 509.

[67] K. Fabel, G. Kempermann, NeuroMolecular Med. 2008, 10, 59.

[68] K. I. Erickson, A. G. Gildengers, M. A. Butters, Dialogues Clin. Neurosci. 2013, 15, 99.

[69] D. Weziak-Bialowolska, P. Bialowolski, P. L. Sacco, Front. Public Health 2023, 11, 1117822.

[70] D. Weber, Age Ageing 2016, 45, 441.

[71] P. National Research Council (US) Committee on Aging Frontiers in SocialPsychology, L. L. Carstensen, C. R. Hartel, in Im 64, National Academies Press (US), 2006.

[72] E. Cohn-Schwartz, R. Khalaila, Healthcare 2022, 10, 2279.

[73] B. D. James, R. S. Wilson, L. L. Barnes, D. A. Bennett, J. Int. Neuropsychol. Soc. JINS 2011, 17, 998.

[74] L. F. Berkman, Lancet 2000, 355, 1291.

[75] L. Fratiglioni, S. Paillard-Borg, B. Winblad, Lancet Neurol. 2004, 3, 343.

[76] N. Scarmeas, Y. Stern, J. Clin. Exp. Neuropsychol. 2003, 25, 625.

[77] A. Ihle, D. Fagot, F. Vallet, N. Ballhausen, N. Mella, M. Baeriswyl, J. Sauter, M. Oris, J. Maurer, M. Kliegel, Neuropsychology 2019, 33, 234.

[78] G. S. Borgeest, R. N. Henson, M. Shafto, D. Samu, R. A. Kievit, PLoS ONE 2020, 15, e0230077.

[79] H. Dejakaisaya, W. Mahikul, N. Na-ek, C. Hirunpattarasilp, PLOS One 2025, 20, e0323863.

[80] H. Takeuchi, R. Kawashima, Front. Neurosci. 2023, 17, 984919.

[81] R. J. Dougherty, T. Hoang, L. J. Launer, D. R. Jacobs, S. Sidney, K. Yaffe, Brain Imaging Behav. 2022, 16, 637.

[82] P. A. Tun, M. E. Lachman, Psychol. Aging 2010, 25, 560.

[83] H. Jiao, Z. Guo, J. Sun, K. Wang, J. Yang, Front. Public Health 2025, 13, 1510418.

[84] N. Charness, W. R. Boot, Curr. Dir. Psychol. Sci. 2009, 18, 253.

[85] S. Sabia, V. T. van Hees, M. J. Shipley, M. I. Trenell, G. Hagger-Johnson, A. Elbaz, M. Kivimaki, A. Singh-Manoux, Am. J. Epidemiol. 2014, 179, 781.

[86] S. M. Dyrstad, B. H. Hansen, I. M. Holme, S. A. Anderssen, Med. Sci. Sports Exerc. 2014, 46, 99.

[87] V. Segura-Jiménez, I. C. Álvarez-Gallardo, A. Romero-Zurita, D. Camiletti-Moirón, D. Munguía-Izquierdo, A. Carbonell-Baeza, J. R. Ruiz, Arch. Phys. Med. Rehabil. 2014, 95, 1903.

[88] S. A. Prince, K. B. Adamo, M. E. Hamel, J. Hardt, S. C. Gorber, M. Tremblay, Int. J. Behav. Nutr. Phys. Act. 2008, 5, 56.

[89] F. Herbolsheimer, M. W. Riepe, R. Peter, BMC Geriatr. 2018, 18, 56.

[90] M. R. Munafò, K. Tilling, A. E. Taylor, D. M. Evans, G. Davey Smith, Int. J. Epidemiol. 2018, 47, 226.

[91] L. C. de Goeij, O. van de Rest, E. J. M. Feskens, L. C. P. G. M. de Groot, E. M. Brouwer-Brolsma, Nutrients 2020, 12, DOI 10.3390/nu12020468.

[92] M. Zhang, X. Dong, Z. Huang, X. Li, Y. Zhao, Y. Wang, H. Zhu, A. Fang, E. L. Giovannucci, Adv. Nutr. 2023, 14, 1170.

[93] H. Kim, Y. Osuka, N. Kojima, H. Sasai, K. Nakamura, C. Oba, M. Sasaki, T. Suzuki, Nutrients 2023, 15, 3181.

[94] T. Suzuki, Y. Osuka, N. Kojima, H. Sasai, K. Nakamura, C. Oba, M. Sasaki, H. Kim, Nutrients 2024, 16, 2800.

[95] Y. Giraldo-Castrillon, J. D. Mendoza, M. Lopez-Cabrera, J. P. Lopez-Lopez, P. Lopez-Jaramillo, Front. Aging 2026, 7, DOI 10.3389/fragi.2026.1709281.

[96] Y. Du, Y. Borné, J. Samuelsson, I. Glans, X. Hu, K. Nägga, S. Palmqvist, O. Hansson, E. Sonestedt, Neurology 2026, 106, e214343.

[97] J. Ni, S. K. Nishi, N. Babio, M. A. Martínez-González, D. Corella, O. Castañer, J. A. Martínez, Á. M. Alonso-Gómez, E. Gómez-Gracia, J. Vioque, D. Romaguera, J. López-Miranda, R. Estruch, F. J. Tinahones, J. Lapetra, J. L. Serra-Majem, A. Bueno-Cavanillas, J. A. Tur, V. Martín-Sánchez, X. Pintó, J. J. Gaforio, A. Barabash Bustelo, J. Vidal, C. Vázquez, L. Daimiel, E. Ros, E. Toledo, O. Coltell, C. Gómez-Martínez, M. D. Zomeño, C. Donat-Vargas, L. Goicolea-Güemez, C. Bouzas, M. Garcia-de-la-Hera, A. Chaplin, A. Garcia-Rios, R. Casas, I. Cornejo-Pareja, J. M. Santos-Lozano, T. Rognoni, C. Saiz, I. Paz-Graniel, M. Malcampo, A. Sánchez-Villegas, I. Salaverria-Lete, A. García-Arellano, H. Schröder, J. Salas-Salvadó, Mol. Nutr. Food Res. 2022, 66, 2101058.

[98] F. Villoz, T. Filippini, N. Ortega, D. Kopp-Heim, T. Voortman, M. R. Blum, C. Del Giovane, M. Vinceti, N. Rodondi, P. O. Chocano-Bedoya, Adv. Nutr. 2024, 15, 100160.

[99] B. S. Klinedinst, S. T. Le, B. Larsen, C. Pappas, N. J. Hoth, A. Pollpeter, Q. Wang, Y. Wang, S. Yu, L. Wang, K. Allenspach, J. P. Mochel, D. A. Bennett, A. A. Willette, J. Alzheimers Dis. JAD 2020, 78, 1245.

[100] A.-J. Tessier, N. Presse, E. Rahme, G. Ferland, L. Bherer, S. Chevalier, J. Gerontol. A. Biol. Sci. Med. Sci. 2021, 76, 2223.

[101] L. Wu, D. Sun, Nutrients 2016, 8, DOI 10.3390/nu8120824.

[102] M. Dehghan, A. Mente, S. Rangarajan, P. Sheridan, V. Mohan, R. Iqbal, R. Gupta, S. Lear, E. Wentzel-Viljoen, A. Avezum, P. Lopez-Jaramillo, P. Mony, R. P. Varma, R. Kumar, J. Chifamba, K. F. Alhabib, N. Mohammadifard, A. Oguz, F. Lanas, D. Rozanska, K. B. Bostrom, K. Yusoff, L. P. Tsolkile, A. Dans, A. Yusufali, A. Orlandini, P. Poirier, R. Khatib, B. Hu, L. Wei, L. Yin, A. Deeraili, K. Yeates, R. Yusuf, N. Ismail, D. Mozaffarian, K. Teo, S. S. Anand, S. Yusuf, The Lancet 2018, 392, 2288.

[103] M. Ozawa, T. Ohara, T. Ninomiya, J. Hata, D. Yoshida, N. Mukai, M. Nagata, K. Uchida, T. Shirota, T. Kitazono, Y. Kiyohara, J. Am. Geriatr. Soc. 2014, 62, 1224.

[104] M. C. Cornelis, S. Weintraub, M. C. Morris, Nutrients 2020, 12, 1969.

[105] M. Barbagallo, A. Springer, C. Vanetta, M. Allemann, P. Lee, S. Saeedi, S. Aeschbacher, M. Luciani, L. H. Bonati, G. Moschovitis, V. Scheu, J. Rutishauser, R. Kobza, M. Di Valentino, P. B. Meyre, N. Rodondi, D. Conen, M. Kühne, S. Osswald, J. H. Beer, J. Am. Heart Assoc. Cardiovasc. Cerebrovasc. Dis. 2024, 14, e034365.

[106] B. Wang, T. Ma, L. Yang, S. He, J. Li, X. Sun, Sci. Rep. 2024, 14, 21063.

[107] Y. Zhang, Y. Liu, Y. Li, Y. Li, X. Gu, J. H. Kang, A. H. Eliassen, M. Wang, E. B. Rimm, W. C. Willett, F. B. Hu, M. J. Stampfer, D. D. Wang, JAMA 2026, e2527259.

[108] M. H. Eskelinen, M. Kivipelto, J. Alzheimers Dis. JAD 2010, 20 Suppl 1, S167.

[109] I. S. Nila, V. M. Villagra Moran, Z. A. Khan, Y. Hong, J. Lifestyle Med. 2023, 13, 83.

[110] J.-S. Jeon, H.-T. Kim, I.-H. Jeong, S.-R. Hong, M.-S. Oh, M.-H. Yoon, J.-H. Shim, J. H. Jeong, A. M. Abd El-Aty, J. Adv. Res. 2019, 17, 85.

[111] D. Restuccia, U. G. Spizzirri, O. I. Parisi, G. Cirillo, N. Picci, Food Chem. 2015, 175, 143.

[112] J. Geertsema, M. Kratochvil, R. González-Domínguez, S. Lefèvre-Arbogast, D. Y. Low, A. Du Preez, H. Lee, M. Urpi-Sarda, A. Sánchez-Pla, L. Aigner, C. Samieri, C. Andres-Lacueva, C. Manach, S. Thuret, P. J. Lucassen, A. Korosi, Neurobiol. Stress 2024, 31, 100641.

[113] K. Ishida, M. Yamamoto, K. Misawa, H. Nishimura, K. Misawa, N. Ota, A. Shimotoyodome, Neurosci. Res. 2020, 154, 35.

[114] M. Kolahdouzan, M. J. Hamadeh, CNS Neurosci. Ther. 2017, 23, 272.

[115] B. Sharma, G. Agriantonis, S. Dawson-Moroz, R. Brown, W. Simon, D. Ebelle, J. Chapelet, A. Cardona, A. Soni, M. Siddiqui, B. Patel, S. Cheerasarn, J. Chang, L. Cobb, F. John, M. M. Hasan, C. Garcia, Z. Shaefee, K. Twelker, N. D. Bhatia, J. Whittington, Nutrients 2025, 17, 1925.

[116] Z. Zhong, H. Dong, S. Zhou, C. Lin, P. Huang, X. Li, J. Zhang, J. Xie, Y. Wu, P. Li, CNS Neurosci. Ther. 2024, 30, e70134.

[117] A. Winiarska-Mieczan, K. Jachimowicz, S. Kislova, M. Kwiecień, Z. Zasadna, D. Yanovych, Biol. Trace Elem. Res. 2023, 201, 425.

[118] G. Strocchi, P. Rubiolo, C. Cordero, C. Bicchi, E. Liberto, Food Chem. 2022, 393, 133406.

[119] H.-J. Kim, S. Cho, D. R. Jacobs, K. Park, Diabetes Res. Clin. Pract. 2014, 106, 145.

[120] M.-K. Shin, K.-N. Kim, J. Korean Med. Sci. 2024, 39, e1.

[121] Y. Wei, Z. Li, H. Lai, P. Lu, B. Zhang, L. Song, L. Zhang, M. Shen, Nutrients 2023, 15, 1354.

[122] L. Morrow, B. Greenwald, Gastroenterol. Nurs. Off. J. Soc. Gastroenterol. Nurses Assoc. 2024, 47, 260.

[123] F. Pulido Valente, Eur. J. Public Health 2023, 33, ckad160.1651.

[124] R. Daviet, G. Aydogan, K. Jagannathan, N. Spilka, P. D. Koellinger, H. R. Kranzler, G. Nave, R. R. Wetherill, Nat. Commun. 2022, 13, 1175.

[125] A. Topiwala, K. P. Ebmeier, T. Maullin-Sapey, T. E. Nichols, NeuroImage Clin. 2022, 35, 103066.

[126] K. M. Gillespie, E. Kemps, M. J. White, S. E. Bartlett, Front. Aging Neurosci. 2025, 17, DOI 10.3389/fnagi.2025.1569069.

[127] H. Essa, H. M. Ali, P. H. Min, D. N. Ali, V. Lowe, R. C. Petersen, D. S. Knopman, E. S. Lundt, C. T. Mester, N. L. Bormann, D.-S. Choi, Alcohol Alcohol 2024, 60, agae080.

[128] G. E. Bond, R. L. Burr, S. M. McCurry, M. M. Rice, A. R. Borenstein, E. B. Larson, Int. Psychogeriatr. 2005, 17, 653.

[129] Y. Akagi, M. Kabayama, Y. Gondo, Y. Masui, S. Yasumoto, N. Klinpudtan, W. Srithumsuk, K. Godai, K. Ikebe, H. Akasaka, S. Yokoyama, Y. Nozato, Y. Takami, Y. Takeya, K. Yamamoto, K. Sugimoto, Y. Arai, H. Inagaki, T. Ishizaki, H. Rakugi, K. Kamide, BMC Geriatr. 2022, 22, 158.

[130] E. T. Reas, G. A. Laughlin, D. Kritz-Silverstein, E. Barrett-Connor, L. K. McEvoy, J. Prev. Alzheimers Dis. 2016, 3, 105.

[131] P. Horvat, M. Richards, R. Kubinova, A. Pajak, S. Malyutina, S. Shishkin, H. Pikhart, A. Peasey, M. G. Marmot, A. Singh-Manoux, M. Bobak, Neurology 2015, 84, 287.

[132] I. Buianova, M. Silvestrin, J. D. Deng, N. Pat, eLife 2026, 14, RP108109.

[133] G. Piumatti, S. C. Moore, D. M. Berridge, C. Sarkar, J. Gallacher, J. Public Health Oxf. Engl. 2018, 40, 304.

[134] S. Kim, Y. Kim, S. M. Park, 2016, DOI 10.1136/bmjopen-2015-010494.

[135] M. C. Reid, P. H. Van Ness, K. A. Hawkins, V. Towle, J. Concato, Z. Guo, J. Geriatr. Psychiatry Neurol. 2006, 19, 98.

[136] M. Zarezadeh, M. Mahmoudinezhad, A. H. Faghfouri, N. Mohammadzadeh Honarvar, Q. R. Regestein, S. I. Papatheodorou, R. A. Mekary, W. C. Willett, Ageing Res. Rev. 2024, 100, 102419.

[137] A. Topiwala, K. P. Ebmeier, Evid. Based Ment. Health 2018, 21, 12.

[138] N. Druffner, D. Egan, S. Ramamurthy, J. O’Brien, A. F. Davis, J. Jack, D. Symester, K. Thomas, J. M. Palka, V. J. Thakkar, E. S. Brown, Alcohol Alcohol 2024, 59, agae035.

[139] M. Cerdá, V. D. Johnson-Lawrence, S. Galea, Soc. Sci. Med. 2011, 73, 1178.

[140] C. K. Lui, W. C. Kerr, N. Mulia, Y. Ye, Drug Alcohol Depend. 2018, 186, 36.

[141] K. H. Beck, F. Zanjani, H. K. Allen, Transp. Res. Part F Traffic Psychol. Behav. 2019, 64, 161.

[142] S. Kim, S. L. Spilman, D. H. Liao, P. Sacco, A. A. Moore, Aging Ment. Health 2018, 22, 550.

[143] H. Molla, T. O’Neill, E. Hahn, R. Lee, H. de Wit, Psychopharmacology (Berl.) 2025, 242, 629.

[144] K. R. Krueger, R. S. Wilson, J. M. Kamenetsky, L. L. Barnes, J. L. Bienias, D. A. Bennett, Exp. Aging Res. 2009, 35, 45.

[145] J. Migeot, M. Calivar, H. Granchetti, A. Ibáñez, S. Fittipaldi, Sci. Rep. 2022, 12, 6048.

[146] K. R. Krueger, P. Desai, T. Beck, L. L. Barnes, J. Bond, C. DeCarli, N. T. Aggarwal, D. A. Evans, K. B. Rajan, JAMA Netw. Open 2025, 8, e2461208.

[147] B. Tari, M. Künzi, V. Raymont, Front. Psychol. 2025, 16, 1542282.

[148] K. Komulainen, C. Hakulinen, J. Lipsanen, T. Partonen, L. Pulkki-Råback, M. Kähönen, M. Virtanen, R. Ruuhela, O. Raitakari, S. Rovio, M. Elovainio, Sci. Rep. 2022, 12, 20818.

[149] L.-Z. Ma, Y.-H. Ma, Y.-N. Ou, S.-D. Chen, L. Yang, Q. Dong, W. Cheng, L. Tan, J.-T. Yu, BMC Med. 2022, 20, 132.

[150] H. Li, F. Cui, T. Wang, W. Wang, D. Zhang, Sci. Rep. 2024, 14, 10313.

[151] L. G. Ioannou, L. Tsoutsoubi, K. Mantzios, G. Gkikas, J. F. Piil, P. C. Dinas, S. R. Notley, G. P. Kenny, L. Nybo, A. D. Flouris, Int. J. Environ. Res. Public. Health 2021, 18, 7698.

[152] J. F. Piil, L. Christiansen, N. B. Morris, C. J. Mikkelsen, L. G. Ioannou, A. D. Flouris, J. Lundbye-Jensen, L. Nybo, Sci. Rep. 2020, 10, 7812.

[153] A. R. Bain, L. Nybo, P. N. Ainslie, Compr. Physiol. 2015, 5, 1345.

[154] L. Nybo, N. H. Secher, B. Nielsen, J. Physiol. 2002, 545, 697.

[155] W. You, Health Sci. Rep. 2025, 8, e70302.

[156] E. A. Kiyatkin, Temp. Multidiscip. Biomed. J. 2019, 6, 271.

[157] K. Kokubun, K. Nemoto, Y. Yamamoto, A. Mitera, Y. Yamakawa, Brain Sci. 2025, 15, 1324.

[158] H. Wang, B. Wang, K. P. Normoyle, K. Jackson, K. Spitler, M. F. Sharrock, C. M. Miller, C. Best, D. Llano, R. Du, Front. Neurosci. 2014, 8, 307.

[159] A. R. Bain, R. L. Hoiland, J. Donnelly, D. Nowak-Flück, M. Sekhon, M. M. Tymko, J. J. Greiner, C. A. DeSouza, P. N. Ainslie, J. Physiol. 2020, 598, 943.

[160] K.-L. Lee, K.-C. Niu, M.-T. Lin, C.-S. Niu, J. Formos. Med. Assoc. 2013, 112, 454.

[161] K. Yoneda, S. Hosomi, H. Ito, Y. Togami, S. Oda, H. Matsumoto, J. Shimazaki, H. Ogura, J. Oda, Front. Neurosci. 2024, 18, DOI 10.3389/fnins.2024.1437216.

[162] R. M. Slominski, J. Y. Chen, C. Raman, A. T. Slominski, Proc. Natl. Acad. Sci. 2024, 121, e2308374121.

[163] K.-N. Yoon, S. Y. Kim, J. Ji, Y. Cui, Q.-L. Quan, G. Park, J.-H. Oh, J. S. Lee, J.-Y. An, J. H. Chung, Y.-S. Lee, D. H. Lee, Exp. Mol. Med. 2024, 56, 1401.

[164] K.-N. Yoon, Y. Kim, Y. Cui, J. Ji, G. Park, J. H. Chung, Y.-S. Lee, J.-Y. An, D. H. Lee, Mol. Brain 2022, 15, 102.

[165] B. Kröger, H.-X. Wang, Ö. Ekblom, J. Wu, H. Westerlund, M. Kivimäki, R. Wang, Commun. Med. 2025, 5, 389.

[166] M. P. Jimenez, N. V. DeVille, E. G. Elliott, J. E. Schiff, G. E. Wilt, J. E. Hart, P. James, Int. J. Environ. Res. Public. Health 2021, 18, 4790.

[167] A. W. Bailey, H.-K. Kang, Int. J. Environ. Res. Public. Health 2022, 19, 16638.

[168] R. S. Geary, D. A. Thompson, J. K. Garrett, A. Mizen, F. M. Rowney, J. Song, M. P. White, R. Lovell, A. Watkins, R. A. Lyons, S. Williams, G. Stratton, A. Akbari, S. C. Parker, M. J. Nieuwenhuijsen, J. White, B. W. Wheeler, R. Fry, D. Tsimpida, S. E. Rodgers, in Green–Blue Space Expo. Chang. Impact Individ.-Level Well-Ment. Health Popul.-Wide Dyn. Longitud. Panel Study Linked Surv. Data, National Institute For Health And Care Research, 2023.

[169] S. J. Geiger, M. P. White, S. M. C. Davison, L. Zhang, O. McMeel, P. Kellett, L. E. Fleming, Commun. Earth Environ. 2023, 4, 166.

[170] E. Cerin, A. P. Matison, M. A. Molina, R. Schroers, W. Li, L. D. Knibbs, V. S. Catts, Y. Wu, M. V. Soloveva, K. J. Anstey, S. Mavoa, G. Poudel, B. Jalaludin, N. A. Kochan, H. Brodaty, P. S. Sachdev, Alzheimers Dement. 2025, 21, e70569.

[171] B. Tari, M. Künzi, C. P. Pflanz, V. Raymont, S. Bauermeister, Front. Public Health 2023, 11, 1244306.

[172] A. Fry, T. J. Littlejohns, C. Sudlow, N. Doherty, L. Adamska, T. Sprosen, R. Collins, N. E. Allen, Am. J. Epidemiol. 2017, 186, 1026.

[173] D. M. Lyall, T. Quinn, L. M. Lyall, J. Ward, J. J. Anderson, D. J. Smith, W. Stewart, R. J. Strawbridge, M. E. S. Bailey, B. Cullen, Brain Commun. 2022, 4, fcac119.

[174] V. Bradley, T. E. Nichols, 2022.

[175] T. Schoeler, D. Speed, E. Porcu, N. Pirastu, J.-B. Pingault, Z. Kutalik, Nat. Hum. Behav. 2023, 7, 1216.

[176] S. van Alten, B. W. Domingue, J. Faul, T. Galama, A. T. Marees, Int. J. Epidemiol. 2024, 53, dyae054.

[177] N. Pat, Y. Wang, R. Anney, L. Riglin, A. Thapar, A. Stringaris, Hum. Brain Mapp. 2022, 43, 5520.

[178] E. Jaqua, E. Biddy, C. Moore, G. Browne, Cureus n.d., 15, e34605.

[179] D. Margină, A. Ungurianu, C. Purdel, D. Tsoukalas, E. Sarandi, M. Thanasoula, F. Tekos, R. Mesnage, D. Kouretas, A. Tsatsakis, Int. J. Environ. Res. Public. Health 2020, 17, 4135.

[180] N. C. B. S. Silva, C. K. Barha, K. I. Erickson, A. F. Kramer, T. Liu-Ambrose, Trends Neurosci. 2024, 47, 402.

[181] D. Fabian, K. M. Kufel, M. Teofilak, N. M. Smyl, O. Śpiołek, J. Siwiec, A. Słowikowska, J. Szatkowska, M. P. Sztyler-Krąkowska, A. A. Wąsowicz, F. Kędziora, Qual. Sport 2025, 37, 57569.

[182] E. Kip, L. C. Parr-Brownlie, Front. Neurosci. 2023, 17, DOI 10.3389/fnins.2023.1092537.

[183] A. Pacholko, C. Iadecola, Hypertension 2024, 81, 991.

[184] D.-Y. Seo, J.-W. Heo, J. R. Ko, H.-B. Kwak, Int. Neurourol. J. 2019, 23, S82.

[185] H. Hu, L. Meng, Y.-L. Bi, W. Zhang, W. Xu, X.-N. Shen, Y.-N. Ou, Y.-H. Ma, Q. Dong, L. Tan, J.-T. Yu, Alzheimers Dement. J. Alzheimers Assoc. 2022, 18, 53.

[186] D. Kellar, S. Craft, Lancet Neurol. 2020, 19, 758.

[187] C. Phillips, Neural Plast. 2017, 2017, 3589271.

[188] Y. Li, B. J. Sahakian, J. Kang, C. Langley, W. Zhang, C. Xie, S. Xiang, J. Yu, W. Cheng, J. Feng, Nat. Aging 2022, 2, 425.

[189] K. Colyer-Patel, L. Kuhns, A. Weidema, H. Lesscher, J. Cousijn, Neurosci. Biobehav. Rev. 2023, 146, 105038.

[190] H. Wright, R. A. Jenks, Age Ageing 2016, 45, 313.

[191] L. J. Dominguez, N. Veronese, L. Vernuccio, G. Catanese, F. Inzerillo, G. Salemi, M. Barbagallo, Nutrients 2021, 13, DOI 10.3390/nu13114080.

[192] S. Song, Y. Stern, Y. Gu, Ageing Res. Rev. 2022, 74, 101551.

[193] H. Hu, Y. Zhao, D. Guo, Y. Deng, H. Luo, Y. Hao, C. Sun, K. Yu, J. Nutr. Health Aging 2025, 29, 100487.

[194] K. M. Keyes, D. Westreich, The Lancet 2019, 393, 1297.

[195] A. Fry, T. J. Littlejohns, C. Sudlow, N. Doherty, N. E. Allen, J Epidemiol Community Health 2016, 70, A26.

[196] A. Tetereva, J. Li, J. D. Deng, A. Stringaris, N. Pat, NeuroImage 2022, 263, 119588.

[197] A. R. Hariri, S. Y. Bookheimer, J. C. Mazziotta, Neuroreport 2000, 11, 43.

[198] W. K. Kirchner, J. Exp. Psychol. 1958, 55, 352.

[199] A. Tetereva, A. R. Knodt, T. R. Melzer, W. van der Vliet, B. Gibson, A. R. Hariri, E. T. Whitman, J. Li, F. Lal Khakpoor, J. Deng, D. Ireland, S. Ramrakha, N. Pat, PNAS Nexus 2025, 4, pgaf175.

[200] A. Tetereva, N. Pat, eLife 2023, 12, DOI 10.7554/eLife.87297.1.

[201] R. C. Gershon, M. V. Wagster, H. C. Hendrie, N. A. Fox, K. F. Cook, C. J. Nowinski, Neurology 2013, 80, S2.

[202] D. E. Hartman, Appl. Neuropsychol. 2009, 16, 85.

[203] D. L. Jackson, J. A. Gillaspy, R. Purc-Stephenson, Psychol. Methods 2009, 14, 6.

[204] W. D. Penny, J. Mattout, N. J. Trujillo-Barreto, in Stat. Parametr. Mapp. Anal. Funct. Brain Images, Academic Press, Ltd, 2007, pp. 454–467.

[205] D. M. Lyall, B. Cullen, M. Allerhand, D. J. Smith, D. Mackay, J. Evans, J. Anderson, C. Fawns-Ritchie, A. M. McIntosh, I. J. Deary, J. P. Pell, PLOS ONE 2016, 11, e0154222.

[206] C. Fawns-Ritchie, I. J. Deary, PLoS ONE 2020, 15, e0231627.

[207] C. L. Craig, A. L. Marshall, M. Sjöström, A. E. Bauman, M. L. Booth, B. E. Ainsworth, M. Pratt, U. Ekelund, A. Yngve, J. F. Sallis, P. Oja, Med. Sci. Sports Exerc. 2003, 35, 1381.

[208] B. E. Ainsworth, W. L. Haskell, S. D. Herrmann, N. Meckes, D. R. Bassett, C. Tudor-Locke, J. L. Greer, J. Vezina, M. C. Whitt-Glover, A. S. Leon, Med. Sci. Sports Exerc. 2011, 43, 1575.

[209] R. Walmsley, S. Chan, K. Smith-Byrne, R. Ramakrishnan, M. Woodward, K. Rahimi, T. Dwyer, D. Bennett, A. Doherty, Br. J. Sports Med. 2021, 56, 1008.

[210] D. Vienneau, K. de Hoogh, M. J. Bechle, R. Beelen, A. van Donkelaar, R. V. Martin, D. B. Millet, G. Hoek, J. D. Marshall, Environ. Sci. Technol. 2013, 47, 13555.

[211] D. J. Lopez, A. Singh, N. T. Waidyatillake, J. C. Su, D. S. Bui, S. C. Dharmage, C. J. Lodge, A. J. Lowe, Br. J. Dermatol. 2022, 187, 704.

[212] Y. E. Tian, V. Cropley, A. B. Maier, N. T. Lautenschlager, M. Breakspear, A. Zalesky, Nat. Med. 2023, 29, 1221.

[213] F. Alfaro-Almagro, M. Jenkinson, N. K. Bangerter, J. L. R. Andersson, L. Griffanti, G. Douaud, S. N. Sotiropoulos, S. Jbabdi, M. Hernandez-Fernandez, E. Vallee, D. Vidaurre, M. Webster, P. McCarthy, C. Rorden, A. Daducci, D. C. Alexander, H. Zhang, I. Dragonu, P. M. Matthews, K. L. Miller, S. M. Smith, NeuroImage 2018, 166, 400.

[214] K. L. Miller, F. Alfaro-Almagro, N. K. Bangerter, D. L. Thomas, E. Yacoub, J. Xu, A. J. Bartsch, S. Jbabdi, S. N. Sotiropoulos, J. L. Andersson, L. Griffanti, G. Douaud, T. W. Okell, P. Weale, I. Dragonu, S. Garratt, S. Hudson, R. Collins, M. Jenkinson, P. M. Matthews, S. M. Smith, Nat. Neurosci. 2016, 19, 1523.

[215] F. Alfaro-Almagro, P. McCarthy, S. Afyouni, J. L. R. Andersson, M. Bastiani, K. L. Miller, T. E. Nichols, S. M. Smith, NeuroImage 2021, 224, 117002.

[216] S. Mansour L., M. A. Di Biase, R. E. Smith, A. Zalesky, C. Seguin, NeuroImage 2023, 283, 120407.

[217] R. S. Desikan, F. Ségonne, B. Fischl, B. T. Quinn, B. C. Dickerson, D. Blacker, R. L. Buckner, A. M. Dale, R. P. Maguire, B. T. Hyman, M. S. Albert, R. J. Killiany, NeuroImage 2006, 31, 968.

[218] Y. Tian, D. S. Margulies, M. Breakspear, A. Zalesky, Nat. Neurosci. 2020, 23, 1421.

[219] C. Destrieux, B. Fischl, A. Dale, E. Halgren, NeuroImage 2010, 53, 1.

[220] M. F. Glasser, T. S. Coalson, E. C. Robinson, C. D. Hacker, J. Harwell, E. Yacoub, K. Ugurbil, J. Andersson, C. F. Beckmann, M. Jenkinson, S. M. Smith, D. C. Van Essen, Nature 2016, 536, 171.

[221] A. Schaefer, R. Kong, E. M. Gordon, T. O. Laumann, X.-N. Zuo, A. J. Holmes, S. B. Eickhoff, B. T. T. Yeo, Cereb. Cortex 2018, 28, 3095.

[222] B. T. Thomas Yeo, F. M. Krienen, J. Sepulcre, M. R. Sabuncu, D. Lashkari, M. Hollinshead, J. L. Roffman, J. W. Smoller, L. Zöllei, J. R. Polimeni, B. Fisch, H. Liu, R. L. Buckner, J. Neurophysiol. 2011, 106, 1125.

[223] T. Chen, C. Guestrin, 2016, pp. 785–794.

[224] L. Breiman, J. Friedman, R. A. Olshen, C. J. Stone, Classification and Regression Trees, Chapman And Hall/CRC, New York, 2017.

[225] L. Breiman, Mach. Learn. 2001, 45, 5.

[226] M. Schonlau, R. Y. Zou, Stata J. 2020, 20, 3.

[227] Y. Tian, A. Zalesky, NeuroImage 2021, 245, 118648.

[228] S. Chopra, E. Dhamala, C. Lawhead, J. A. Ricard, E. R. Orchard, L. An, P. Chen, N. Wulan, P. Kumar, A. Rubenstein, J. Moses, L. Chen, P. Levi, A. Holmes, K. Aquino, A. Fornito, I. Harpaz-Rotem, L. T. Germine, J. T. Baker, B. T. T. Yeo, A. J. Holmes, Sci. Adv. 2024, 10, eadn1862.

[229] J. Chen, L. Q. R. Ooi, T. W. K. Tan, S. Zhang, J. Li, C. L. Asplund, S. B. Eickhoff, D. Bzdok, A. J. Holmes, B. T. T. Yeo, NeuroImage 2023, 274, 120115.

[230] K. Nimon, M. Lewis, R. Kane, R. M. Haynes, Behav. Res. Methods 2008, 40, 457.

[231] C. Viswesvaran, Pers. Psychol. 1998, 51, 223.

